# Screening & diagnosing errors in longitudinal measures of body size

**DOI:** 10.1101/2020.11.19.20234872

**Authors:** AK Wills

## Abstract

This paper presents a novel multi-step automated algorithm to screen for errors in longitudinal height and weight data and describes the frequency and characteristics of errors in three datasets. It also offers a taxonomy of published cleaning routines from a scoping review.

Illustrative data are from three Norwegian retrospective cohorts containing 87,792 assessments (birth to 14y) from 8,428 children. Each has different data pipelines, quality control and data structure. The algorithm contains 43 steps split into 3 sections; (a) dates, (b) Identifiable data entry errors, (c) biologically impossible/ implausible change, and uses logic checks, and cross-sectional and longitudinal routines. The WHO cross-sectional approach was also applied as a comparison.

Published cleaning routines were taxonomized by their design, the marker used to screen errors, the reference threshold and how threshold was selected. Fully automated error detection was not possible without false positives or reduced sensitivity. Error frequencies in the cohorts were 0.4%, 2.1% and 2.4% of all assessments, and the percentage of children with ≥1 error was 4.1%, 13.4% and 15.3%. In two of the datasets, >2/3s of errors could be classified as inliers (within ±3SD scores). Children with errors had a similar distribution of HT and WT to those without error. The WHO cross-sectional approach lacked sensitivity (range 0-55%), flagged many false positives (range: 7-100%) and biased estimates of overweight and thinness.

Elements of this algorithm may have utility for built-in data entry rules, data harmonisation and sensitivity analyses. The reported error frequencies and structure may also help design more realistic simulation studies to test routines. Multi-step distribution-wide algorithmic approaches are recommended to systematically screen and document the wide range of ways in which errors can occur and to maximise sensitivity for detecting errors, naive cross-sectional trimming as a stand-alone method may do more harm than good.

## Introduction

There is a wealth of large-scale population-based longitudinal data on body size from across the (1) world that has been used in thousands of research articles. While height and weight may seem the most trivial of measures to obtain accurately, errors can arise at multiple points in the measurement and data handling schedule. Further, the data flow and quality assurance strategies implemented are rarely the same between studies or even within a study at different data collection waves, particularly when data are from a mix of primary and secondary sources. Errors are thus unlikely to have the same frequency or structure either between or within a study. This may pose a threat to the validity of research findings and may bias many types of comparison.

Errors from measurement and protocol can be minimised with calibration, standardisation and training, leaving in theory only small random Gaussian-type error. Errors from protocol non-compliance and data handling are more significant. Processes such as double data entry and range delimiters can mitigate these, but they are not always implemented, particularly for routinely collected clinical data, and will not pick up all errors. Data entry error rates have been shown to vary from 0.05 to 9% (2) and since growth or change is characterised by repeated measures, as well as sex and age (often derived from two dates), the error rate per individual will be much higher. One study found at least one error in the clinical weight measurements of 20% of patients (3). A single error will also contaminate measures of growth and any derived variables such as BMI.

Clear reporting of the dataflow, and of error types and frequencies is an important aspect of open and reproducible science (4). It is of relevance for designing better data handling processes and error detection methods, for facilitating between study comparisons, and of importance to multi-cohort and collaborative research that require data harmonisation (5). It has been recommended that reports describe data cleaning methods and summarise error types and frequencies (6, 7), however, compliance is low and where reported, detail is often insufficient (8). Hence, most cleaning still takes place under the hidden degrees of freedom of the researcher (9) and little is known about the characteristics of errors.

The aim of this paper is to describe the frequency and characteristics of errors in the longitudinal height and weight data from three retrospective cohort studies containing primary and secondary data sources. To achieve this, a scoping review (10) of different cleaning approaches was done and an algorithm was developed to screen for errors. Both are reported in this paper and code is available. To provide context, the most commonly used method for identifying biologically implausible values (BIV), the WHO approach (11), was also applied and is compared to the algorithm developed here.

## Methods

### Data

Data are from three retrospective Norwegian cohorts, comprising 8,428 children and 87,792 assessments (age range: birth to 14y). Children were recruited at the time of school health service clinics in year 3 (age 8y) for cohorts 2010 & 2015 (the year the cohort was established), and year 8 (age 13y) for the 2017 cohort.

There are several differences in the data collection and quality control processes (Table 1), and data structure (Table S2, OSM file 1) between cohorts. All cohorts contain routinely collected (secondary) data linked from the national medical birth registry (MBR) and from the national child measurement program (health card), and primary research data from the school clinics. Each data source has different data handling processes, for example, there are two transcription steps from measurement to database in the MBR and school clinic data and three for the health card data. The 2010 dataset had undergone some cleaning (however no code was available and we had no access to the raw data). Some checks had also been done on the school clinic data at age 8y in the 2015 dataset, and both the 2010 and 2015 school clinic data had been double-entered. The only quality control for the 2017 cohort were range delimiters on the electronic data entry form (Table 1).

**Table 1.**
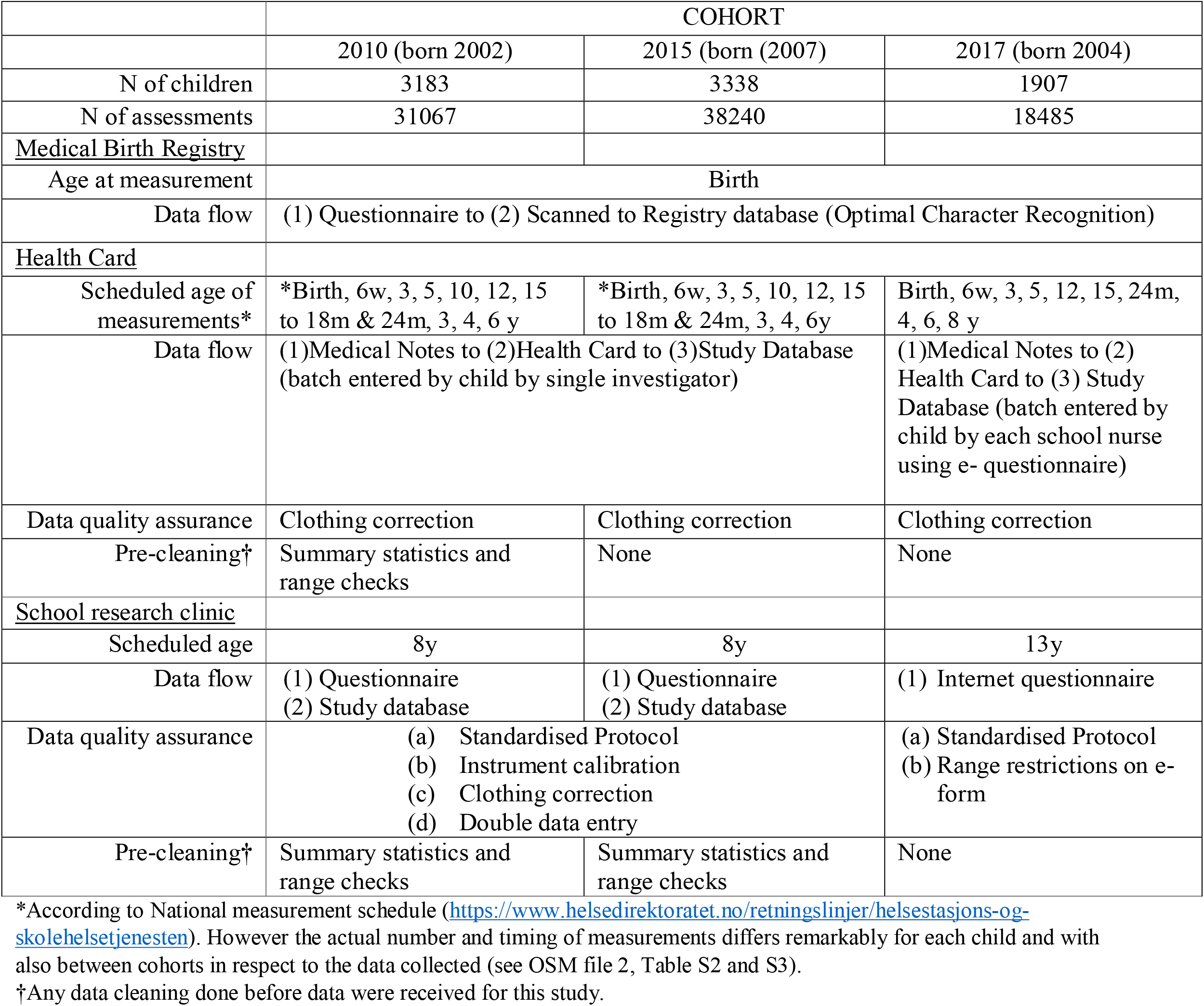
Information about the Cohorts and data source

|  | COHORT |  |  |
| --- | --- | --- | --- |
|  | 2010 (born 2002) | 2015 (born (2007) | 2017 (born 2004) |
| N of children | 3183 | 3338 | 1907 |
| N of assessments | 31067 | 38240 | 18485 |
| <u>Medical Birth Registry</u> |  |  |  |
| Age at measurement | Birth |  |  |
| Data flow | (1) Questionnaire to (2) Scanned to Registry database (Optimal Character Recognition) |  |  |
| <u>Health Card</u> |  |  |  |
| Scheduled age of measurements* | *Birth, 6w, 3, 5, 10, 12, 15 to 18m & 24m, 3, 4, 6 y | *Birth, 6w, 3, 5, 10, 12, 15 to 18m & 24m, 3, 4, 6y | Birth, 6w, 3, 5, 12, 15, 24m, 4, 6, 8 y |
| Data flow | (1)Medical Notes to (2)Health Card to (3)Study Database (batch entered by child by single investigator) |  | (1)Medical Notes to (2) Health Card to (3) Study Database (batch entered by child by each school nurse using e- questionnaire) |
| Data quality assurance | Clothing correction | Clothing correction | Clothing correction |
| Pre-cleaning† | Summary statistics and range checks | None | None |
| <u>School research clinic</u> |  |  |  |
| Scheduled age | 8y | 8y | 13y |
| Data flow | (1) Questionnaire<br>(2) Study database | (1) Questionnaire<br>(2) Study database | (1) Internet questionnaire |
| Data quality assurance | (a) Standardised Protocol<br>(b) Instrument calibration<br>(c) Clothing correction<br>(d) Double data entry |  | (a) Standardised Protocol<br>(b) Range restrictions on e-form |
| Pre-cleaning† | Summary statistics and range checks | Summary statistics and range checks | None |

\*According to National measurement schedule (https://www.helsedirektoratet.no/retningslinjer/helsestasjons-og-skolehelsetjenesten). However the actual number and timing of measurements differs remarkably for each child and with also between cohorts in respect to the data collected (see OSM file 2, Table S2 and S3).
†Any data cleaning done before data were received for this study.

Most measurements were taken between birth and 2y of age (Table S2, OSM file 1). The density of measurements was greatest in the 2015 cohort where all health card measurements were extracted, and most sparse in the 2017 cohort where only one measurement at defined target ages of the national measurement schedule was extracted (figure S2, OSM file 1). Almost one-third of children in the 2015 cohort had >14 measures compared to none in the other two cohorts. Less than 10% of children had <4 measurements, and more than 80% had at least seven (Table S3, OSM file 1). Prior to cleaning, complete duplicate assessments (age, height, weight) and assessments missing age or both height (HT) and weight (WT) were removed. Data were also harmonised to maximise the similarity between and within cohorts (details in OSM file 1).

### Scoping review of approaches for identifying errors

Lawman et al’s., 2016 (8) review of approaches to identify height and weight BIVs was used as a starting to identify the different approaches that have been used for error detection. The 12 studies reported in their Table 1 (11-22) were screened along with all subsequent citations of Lawman et al (8) up to Oct 2020 (n=12) (3, 9, 23-32). The reference lists of these citing papers were also screened to identify methodological papers describing cleaning algorithms or approaches (n=5) (7, 33-36). The citations in these papers were then also screened (n=1) (37) (see Fig S1 in OSM file 1) Table S1 (OSM file 1) offers a taxonomy of the cleaning approaches found based on their design (cross sectional, longitudinal), the marker used to detect error (eg; raw score, standardised score, residual) and how the threshold for error was selected (eg, external or internal reference etc). Variations of these approaches exist and some studies have used a combination of methods which is not reflected in the table.

Cross-sectional trimming of absolute or externally age and sex standardised outliers (eg; WHO z-scores) was the most frequently used method (8) (16) (11, 12). If the threshold set is too low then this approach produces a large number of false positives since errors and true outliers are conflated (94% of outliers were deemed true in one study (15). Similarly, sensitivity can be poor since erroneous inliers are not screened (50% of flagged weight errors were inliers in one dataset (9). Selective removal of errors and true values from the ends of the distribution may also introduce other unknown biases, for example, removing outliers, may link missingness and error in the dataset to the true value. This was seen in a simple descriptive setting where applying this approach caused obesity prevalence to be underestimated (34). Standard statistical methods such as the Dixon test have also been tried but lacked sensitivity even for secondary data (35).

Longitudinal approaches can estimate the internal (within-subject) consistency of repeated values which means the entire distribution is screened and erroneous inliers detected. Comparisons with cross-sectional approaches showed improved sensitivity and specificity (28, 29). Approaches include the use of change scores (13, 18) and % change (3), residuals from OLS regression (28, 32), multilevel models (MLM) (23, 35) and non-parametric smoothing routines (9), conditional growth scores (36) and ratios of Euclidean distances between a set of 3 measures (3). Some studies report using manual verification of growth histories (9, 26) (15) (3) (17), which is considered the gold standard approach. Certain methods require more data points which may be a limitation, and some methods perform poorly when the error load (magnitude and frequency of errors) is high (9, 35).

Logic checks based on data entry error mechanisms are another screening/diagnostic method and can be cross-sectional eg; digit errors (29), or longitudinal eg; measures carried forward (9). These checks are rarely reported, which might reflect their status as a routine part of quality control, but it may also reflect a lack of systematic rigor and transparency in data handling processes. For secondary data, these checks will have more utility and where errors have been described, they comprise the majority of errors. For example, in one study more than 96% of detected WT errors could be attributed to a mechanism (9).

The prevalence of flagged suspicious values or errors (and hence sensitivity and specificity) depends on the threshold cut points. Higher thresholds flag fewer false positives but have lower sensitivity. This selection has been made based on biologically impossible limits, arbitrary statistical-based limits eg; flagging values greater than a nominal number of standard deviations (11), and iteratively eg; through visual inspection of charts containing flagged values (17). One more objective method based on the slope of the cumulative distribution of the error marker was reported (3) but it requires a very large sample size.

With respect to a framework for cleaning, Van den Broeck et al. (2005) (7) described a three-step process. First, suspicious values are flagged (screening), second, flagged values are investigated and classified (diagnosis), and third, errors are corrected or removed (treatment). Some aspects of cleaning can be combined into a single step, eg; impossible values can be screened and diagnosed using the same threshold. However, no single-step automated method for screening and diagnosis was able to detect errors without false positives and harm to sensitivity. These findings are unsurprising given the multitude of ways in which error can enter a dataset. Hence, although single-step automated approaches are simple and quick, the selective deletion of true values and screening of particular types of error may introduce bias and. the effect of measurement error on analyses are complex and difficult to predict (38). It is thus important that cleaning is done in a way that enhances the validity of the substantive analyses and does not introduce additional unknown biases.

### Overview of the cleaning algorithm

#### Principles

The original plan was to implement and optimise a previously published automated cleaning routine. However, based on the literature review and exploratory work implementing some of the published routines it was clear that while a fully automated approach is reproducible, it is not accurate, and removing true values should be avoided particularly if data are used for future unknown analyses. The following principles were thus developed to guide the cleaning approach.

1. *Only errors should be removed (38, 39)*.
2. *Screen the entire distribution of values*
3. *Classify and diagnose errors*.
4. *Automate screening*.
5. *Automate diagnosis of errors as much as possible but manually validate all automated decisions*

With respect to point (1) it was useful to clarify the definitions of the jargon used in the cleaning literature such as *implausible* (Table S4, OSM file 1), which means ‘subjectively highly unlikely’, and to adhere to these in the formulation of error rules. In practice this meant that a value was kept if it’s veracity was uncertain. Point (2) meant favouring longitudinal routines that require fewer data points to run. Point (3) facilitates documentation and error reporting, and the creation of screening rules by defining either the error mechanism, or the way in which the value is impossible/implausible. Point (4) ensures screening is systematic and reproducible. Point (5) ensures cleaning was as accurate as the gold standard of manual inspection, minimises falsely removing true values and allows the accuracy of automated procedures to be estimated by tagging automated decisions that were either over-ridden or could not be automated.

#### Development

The algorithm was developed using information on the data-structure, measurement protocols and data flow, and through an iterative process of data exploration and visualisation. Some errors were detected ad-hoc from growth visualisations, and screening and diagnostic rules subsequently developed and implemented. Sometimes the order of the algorithm was changed to maximise the performance of each screening step in terms of the ratio of false positives to flagged values.

#### Outline of algorithm

The steps of the algorithm are fully outlined in OSM file 2. Briefly, there are 43 steps, structured into 3 sections. At each step, flagged values and their treatment are documented into saved variables. Part (A) cleans dates, part (B) targets errors linked to a data entry error mechanism, for example, switched heights and weights and duplicates, and part (C) screens for errors that cause an impossible or implausible absolute change in size. To maximise sensitivity, after the data had been passed through (A) to (C), the automated routine described by Daymount et al., 2017 (deviations from smoothed SD scores (9)) was run on the entire sample to check for missed longitudinal inconsistencies/errors. Similarly, any missed cross-sectional outliers with an absolute modified SD score (see below) greater than 3 were also checked. Growth histories of both HT and WT containing the flagged values from these routines were manually inspected. If any were deemed errors, thresholds in the respective part of the algorithm corresponding to the type of error were adjusted to ensure they were identified and the algorithm was rerun.

#### Modified SD scores

Several steps involve screening SD scores to flag or diagnose errors. LMS derived z-scores have properties that mean extremely high or low HT or WT values can take relatively unremarkable z-scores; this makes them ill-suited for screening errors. A modified SD score as proposed by CDC (12) was thus used. Here the z-score is expressed relative to half the distance between a z-score of 0 and +2 in raw anthropometric units if the measure is above the age-standardised median or half the distance between a z-score of -2 and 0 if the value is below the median. The closeness that the median z-score tracks to zero across age will also depend on the growth reference used. This can also complicate screening since any threshold will then need to account for age. To remove this age-effect, the modified SD scores are centred at the sample median for each age (9).

#### Longitudinal routines for checking internal consistency of values and diagnosing errors

Four longitudinal approaches/ routines were ultimately used in various screening and diagnostic processes. Briefly, these are (i) internal consistency of modified SD scores; (ii) jackknife residuals; (iii) velocity aberrations of the child’s growth history and (iv) Manual inspection (detailed in OSM file 2). The first three create markers of internal consistency which are then compared to thresholds to screen and/or diagnose errors. MLM-based residuals were tried but performed poorly, partly because their validity depends on a well-specified growth model at the individual level which is not trivial and requires several parameters, and partly because of the inherent shrinkage bias of the best linear unbiased predictors which is dependent on the individual-level fit and number of repeated measures of each child.

#### Thresholds

Several of the steps use thresholds to screen for errors. These were selected iteratively using visual inspection of the growth histories of individuals with flagged values, the objective was to select a threshold that captured all errors in that step with no false positives. If this was not possible, a threshold was selected that captured all errors with the screened characteristic but minimised the number of false positives.

### Analysis

The screening algorithm was described using plots of exemplars to illustrate some of the different steps. Several sets of descriptive statistics were produced to describe the errors. The frequencies and percentages of errors by each screening step and totals across all steps within each cohort were tabulated as was the frequency of errors per child. To understand the accuracy of the automated routines and the proportion of records that required manual screening and cleaning, the proportion of errors that were unable to be automated and the proportion of values where the automated diagnosis was over-ridden (in some cases values were kept ie; false positives) was calculated. Cumulative percentages across the algorithm steps were also calculated and plotted. Spaghetti plots of all individuals before and after cleaning HT and WT were produced. The distributions of HT and WT errors by their age and sex standardised percentile position were plotted and the proportion of errors that were inliers and outliers (|SD|>3) was tabulated.

To explore whether errors were dependent on characteristics of individuals, the distribution of the most contemporary weight and height measure by whether a child had an error or not was plotted. Multilevel logistic models (MLM) were estimated to understand if errors were patterned by county, maternal age, education and parity, and parental origin of birth (information provided by Statistics Norway). These were adjusted for the number of measurements of each child.

As a comparison with the most commonly used cleaning method, the WHO cross-sectional cut-offs (11) for erroneous values were applied to the unclean data and the sensitivity, specificity, positive predictive (PPV) and negative predictive value (NPV) was calculated compared to the algorithm. The prevalence of IOTF overweight, obesity and grade 1 thinness (40) were also calculated in the longitudinal algorithm cleaned dataset and in the dataset after excluding the WHO cross-sectional outliers.

The algorithm was created in Stata (v16.1) and R (v4.03) was used for some visualisations.

## Results

### Description of algorithm

Online Supplementary file 3 contains exemplar plots to illustrate the steps in the algorithm that are described below.

#### Part A Dates

Part A was implemented without access to the anthropometric data on a secure server to comply with the 2018 General Data Protection Regulation guidance and so is limited in scope and not integrated with other parts of the algorithm. Screening was based on comparing the date variables to parameters that define the study’s time-frame (index birth year to end of data collection) and the participant’s time-frame in the study (DoB to date of final assessment). Some errors were corrected under reasonable assumptions, for example, checking each numeric place holder corresponding to millennium, century, decade and year allowed corrections of the sort: 1012 to 2012; 2102 to 2012; 2042 to 2012 and 2001 to 2010. The relative frequency of detected data errors was ∼0.3% in cohort 2015 & 2017 and zero in cohort 2010 (Table 2).

**Table 2.**
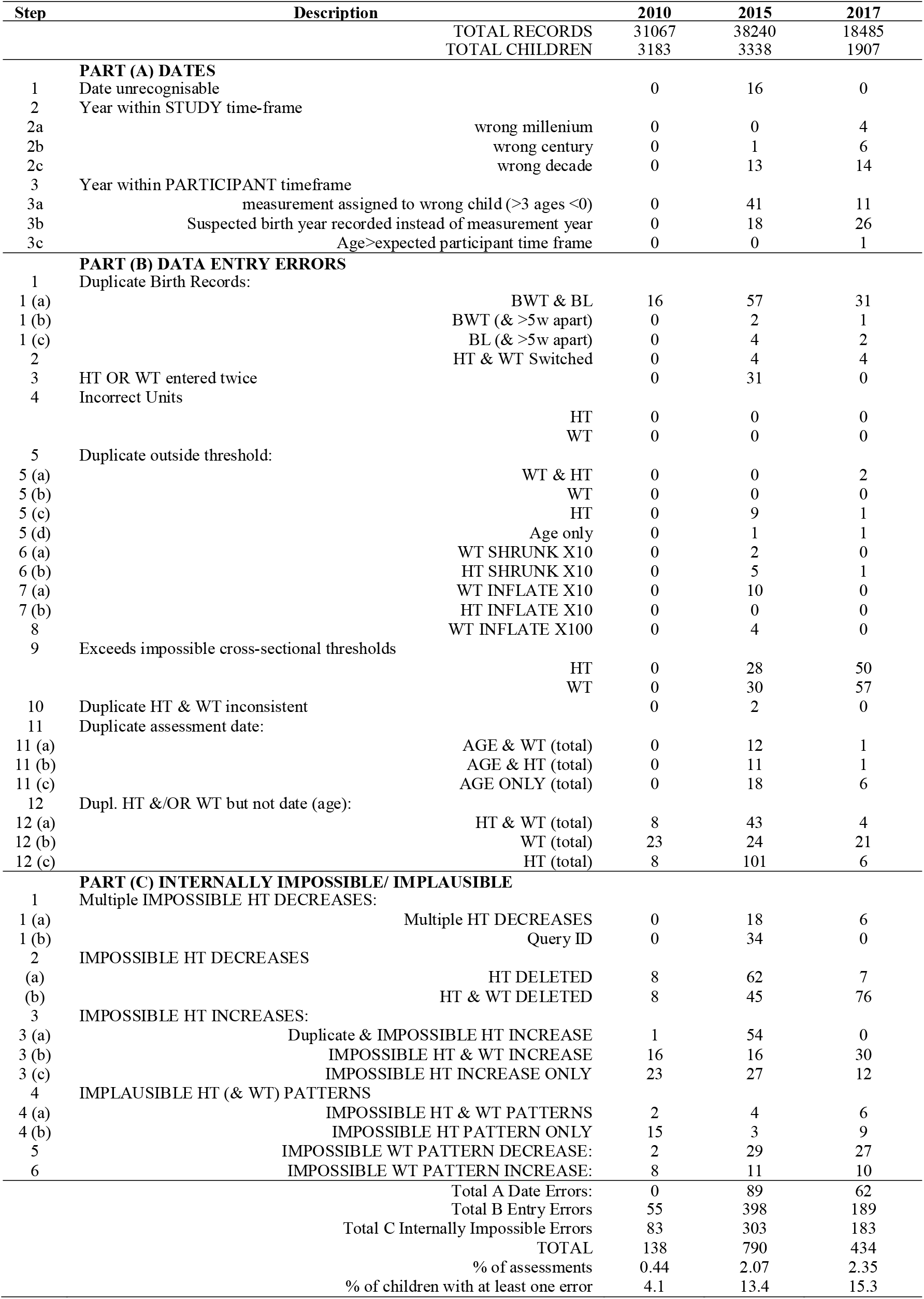
Frequency of errors (n) by each step of the screening algorithm.

#### Part B Identifiable data entry error

Some of the steps in part B use longitudinal methods to automate the error diagnosis of duplicate values and as expected these automated routines performed poorly when the error load was high, particularly for the 2015 dataset (figure 1) which had a high number of duplicates compared to other cohorts (Table S5, OSM file 1). The order of the steps thus partly reflects the need to reduce the error load and many of the first ten steps deal with errors that are large in magnitude and easy to diagnose. Impossible absolute age-related values for size were also removed at this stage (step B9, OSM file 2), Table 2 shows that 15% (60/398) and 57% (107/189) of the errors in part B were picked up in this step for cohorts 2015 and 2017, while cohort 2010 had zero impossible absolute values.

**Figure 1.**
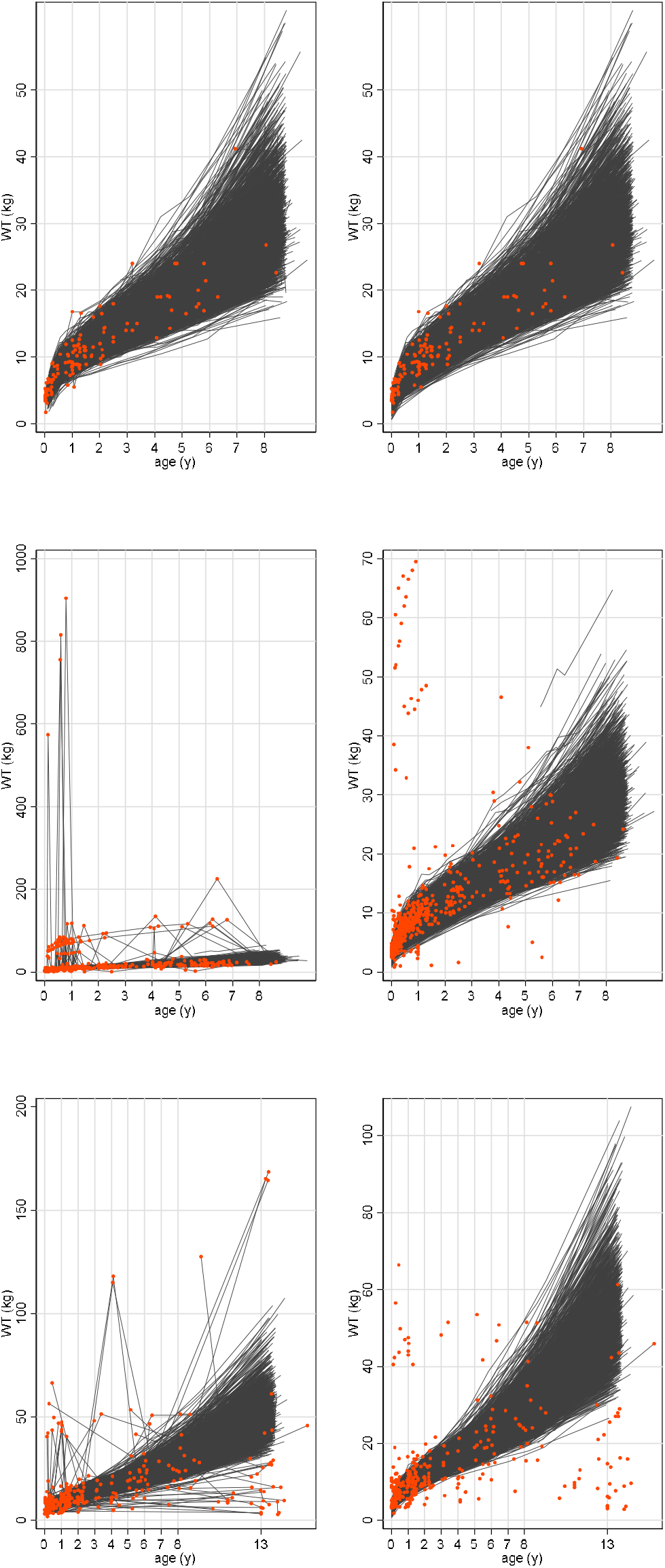
Spaghetti plots of weight before (left column)* and after (right column) cleaning weights and in each of the 3 cohorts, 2010 (top row), 2015 (middle row) 2017 (bottom row). The red points are the diagnosed errors.

Six permutations of duplicate age, HT or WT values are possible when either one or two of the three variables are duplicated (steps B11 & 12). Duplicates of HT or WT (but not both) on the same date cannot be true. On the other hand, duplicates on separate dates are potentially plausible, particularly if only one of HT or WT is duplicated. However, they are implausible if they are a wide age-interval apart and alongside concomitant increases in the other measure. Visualisations were used to set thresholds for these characteristics such that no true values were flagged. Jackknife residuals were used to diagnose the erroneous duplicate where values occurred on the same date or where both HT & WT was duplicated (exemplars shown figure S1, OSM file 3). For duplicate HTs or WTs on separate dates (step B12), an additional criterion was applied based on whether a duplicate caused an aberration to the growth velocity history of the child above an iteratively selected threshold, and the duplicate with the highest velocity aberration was identified as the error (figure S2, OSM file 3).

Despite a substantial time-investment to optimise the automated diagnostic rules and thresholds, on a small number of occasions the automated routine selected the incorrect value as the error and was manually over-ridden (figure 2 & exemplar in figure S3, OSM file 3). Automated selection was also not possible or too time-consuming for individuals that had >1 duplicate, >1 pair of duplicates (high error load), where duplicates were not adjacent in time, and/ or where individuals had ≤3 measurements (figure S4, OSM file 3). In these scenarios, the erroneous duplicate was diagnosed manually. Manual diagnosis was particularly needed in the 2015 dataset for duplicate age and HT values (step B12c) (92/101 errors flagged in this step).

**Figure 2.**
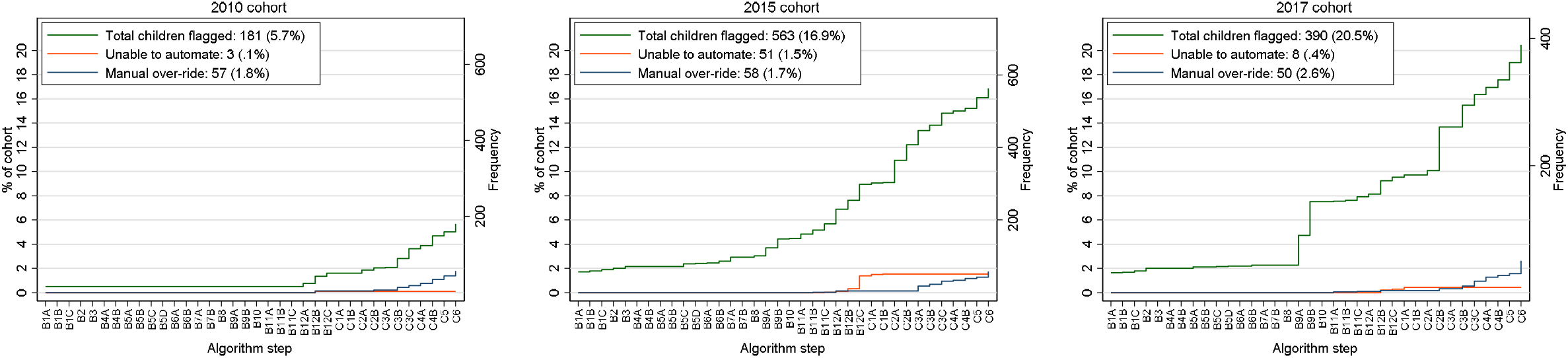
Cumulative percent and frequency of children with a flagged value at each successive step of the HT and WT screening algorithm, also shown is the cumulative percent and frequency of children where automated diagnosis of errors was not possible (orange line) and where the automated diagnostic rule was over-ridden after visualisation (blue line).

#### Part C Impossible and implausible changes and patterns of growth

Impossible HT decreases were first addressed (parts C1 & C2), these often displayed as spikes in the growth history plots. A small number of individuals had >1 height decrease, the high error load for these individuals made it difficult to automate the identification of the erroneous values causing the decreases and so the errors were diagnosed manually. One child’s growth history containing two HT decreases also suggested that values from 2 children had been assigned to a single ID, another possible error mechanism (figure S5, OSM file 3). Jackknife residuals were used to automate the diagnosis of single HT decreases (part C2). Often the HT decrease was accompanied by a decrease in WT, possibly due to an incorrect date, and when this exceeded a threshold, both HT and WT values were flagged as errors (figure S6, OSM file 3).

Part C3 screens impossible absolute HT increases and uses absolute age-related velocity thresholds based on the Tanner and WHO velocity references (41, 42). The thresholds were weighted for each pair of measures by the age period over which the velocity occurred. Additional criteria were set to minimise flagging high velocity values caused by random measurement error from two measurements close in time and to exclude expected pre-term catch-up growth. In the 2015 cohort, many of the flagged impossible HT increases were also duplicates spaced <90 days apart (n=54/97) that weren’t dealt with in part B12 (figure S7, OSM file 3). Likewise, some flagged impossible HT increases were accompanied by suspicious WT increases, which may again reflect a date error (figure S8, OSM file 3). Occasionally the thresholds captured false positive and were over-ridden (exemplars shown in figure S9, OSM file 3).

Impossible/ implausible patterns of height growth were then screened (part C4) using thresholds based on change on the SD scale with lower thresholds set for shorter time intervals (OSM file 2). Unlike the absolute thresholds in parts C1 to C3, these routines tended to captured errors manifesting as implausible change over longer age-intervals (figure S10, OSM file 3). By this stage, errors were ever smaller and more variable in their presentation. This made it more difficult to accurately automate detection and diagnosis as shown by proportion of flagged values where the automated decision was overridden (figure 2). However, even with manual verification, discerning errors from true values among those flagged was not always straightforward. Referring to the definitions of impossible and implausible helped, figure S10 (OSM file 3) shows an example which was not removed because of uncertainty over whether it was an error.

The final parts of the algorithm screen for values that cause impossible WT patterns (figure S11 & S12, OSM file 3). WT trajectories exhibit more variability compared to HT and the distinction between plausible and implausible/impossible was less clear. In the final step, the majority of values flagged from screening were over-ridden and kept (n=48/77).

After passing data through the algorithm the longitudinal SD smoothing and cross-sectional thresholds were applied to check for missed errors. Figures S13-16 (OSM file 3) plot the growth histories in those that were flagged in the final cleaned 2010 dataset. A few of the remaining flagged values were suspicious but not deemed impossible or implausible.

### Automated versus manual cleaning

Figure 2 shows the cumulative percentage of children with flagged values at each successive step of the algorithm, the cumulative percentage where automation was not possible, and the cumulative percentage where the automated diagnosis was over-ridden. The percentage of children flagged from screening was 5.7%, 16.7% and 20.5% in the 2010, 2015 and 2017 datasets respectively, while including only those steps which required manual verification flagged 5.2%; 12.4% and 13.4% of children. In a minority of children, the automated routine for error diagnosis was not possible or not pursued because it was too time-consuming to code (2010: 0.1%; 2015: 1.5%; 2017: 0.4%). Similarly, the automated decision was over-ridden in a small percentage of all children (2010: 1.8%; 2015: 1.7%; 2017: 2.6% of all children).

### Error frequencies

Figure 1 shows a spaghetti plot of the WT data in each cohort before and after removal of detected errors (red points). The differences in the error structure between the cohorts illustrated here is also reflected in the differences between cohorts in the frequency of errors at each cleaning step (Table 2).

Cohort 2010 had the fewest total errors, only 0.4% v 2.1% and 2.4% of assessments in cohorts 2015 and 2017; and the smallest percentage of children with at least one error, 4.1% v 13.4% and 15.3% assessments in cohorts 2010, 2015 and 2017 respectively (Table 2). The majority of children had only one error, only 0.2%, 2.5% & 2.0% of the children in datasets 2010, 2015 and 2017 had more than one error (Table 3).

**Table 3.** Frequency of number of errors per child in each cohort

| Number of errors | 2010 |  | 2015 |  | 2017 |  |
| --- | --- | --- | --- | --- | --- | --- |
|  | Freq | % | Freq | % | Freq | % |
| 0 | 3053 | 95,90 | 2903 | 86,90 | 1640 | 86 |
| 1 | 123 | 3,90 | 351 | 10,50 | 230 | 12,1 |
| 2 | 4 | 0,13 | 43 | 1,30 | 32 | 1,7 |
| 3 | 2 | 0,06 | 19 | 0,57 | 4 | 0,21 |
| 4 |  |  | 11 | 0,33 | 2 | 0,1 |
| 5 |  |  | 4 | 0,12 |  |  |
| 6 |  |  | 6 | 0,18 |  |  |
| 7 |  |  |  |  |  |  |
| 8 |  |  | 1 | 0,03 |  |  |
| ... |  |  |  |  |  |  |
| 36 |  |  | 1 | 0,03 |  |  |
| TOTAL | 3182 | 100 | 3339 | 100 | 1908 | 100 |

### Characteristics of errors

Figure 3 shows the distribution of errors by age and sex standardised percentile. Many errors could be classified as inliers, for example, >2/3 of errors were within limits of ±3 modified SD scores in the 2010 and 2015 cohorts, and just over a 1/3 for the 2017 cohort (Table 4). Figure 4 shows the distribution of the most contemporary measure of child size by whether the child had an error or not. Children with errors had a similar distribution to those without error.

**Table 4.** Frequency (%) of erroneous inliers and outliers* in each cohort

|  | Height |  | Weight |  | Total |  |
| --- | --- | --- | --- | --- | --- | --- |
|  | Inliers | Outliers | Inliers | Outliers | Inliers | Outliers |
| 2010 | 77/105<br>(73%) | 28/105<br>(26.3%) | 73/83<br>(88%) | 10/83<br>(12.1%) | 150/188<br>79.8% | 38/188<br>20.2% |
| 2015 | 360/536<br>(67%) | 176/556<br>(32.8%) | 243/344<br>(70.6) | 101/344<br>29.4% | 603/880<br>(68.5%) | 277/880<br>(31.4%) |
| 2017 | 73/260<br>28.1% | 185/260<br>71.2% | 118/269<br>43.9% | 151/269<br>56.1% | 191/529<br>36.1% | 336/529<br>63.5% |

\*Outlier defined as a modified CDC SD score $>3$ or $<-3$
\*Based on raw data after age had been cleaned (it is not possible to calculate age-standardised z-scores when age is outside the range of the growth reference)

**Figure 3.**
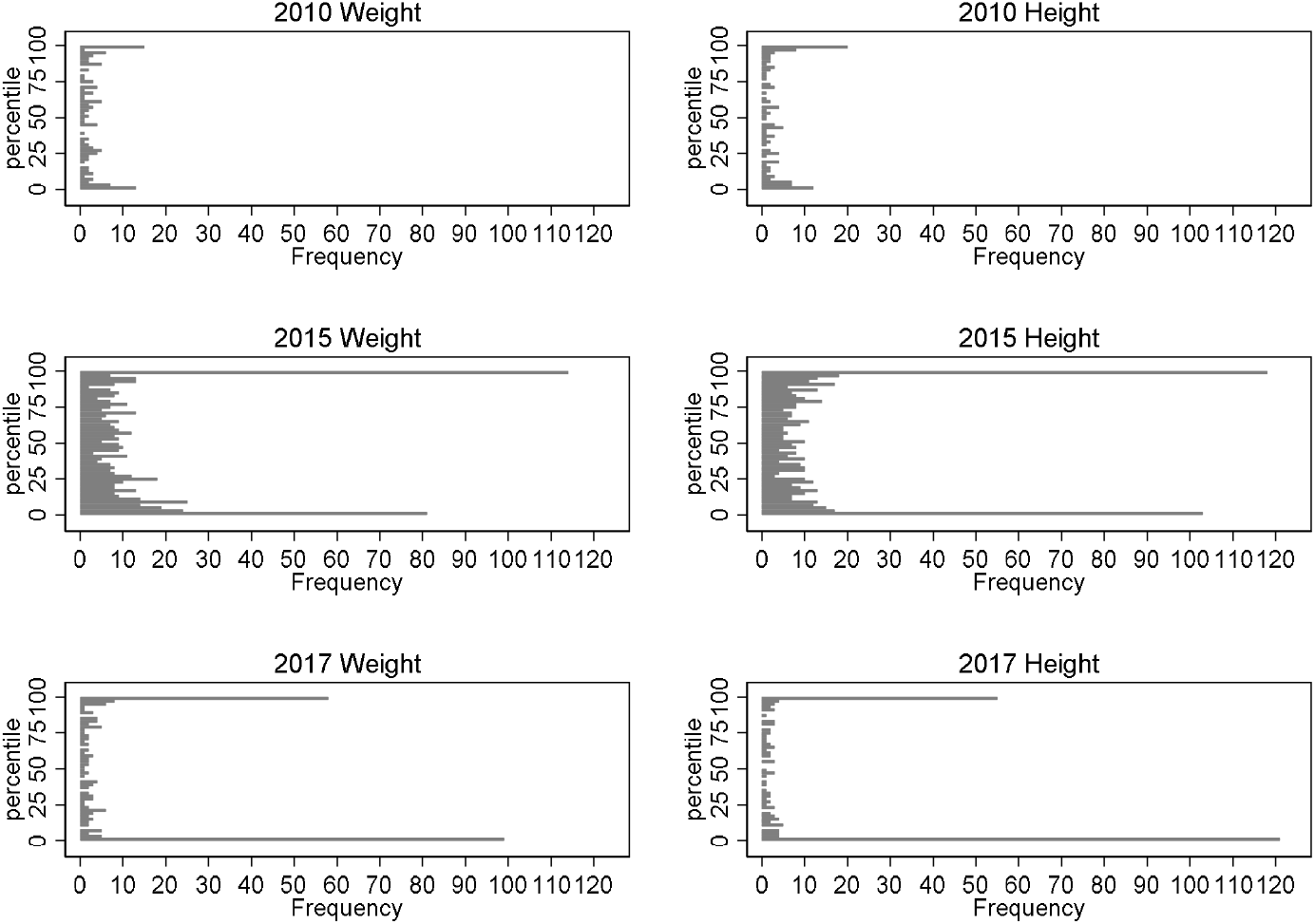
Histogram of errors according to their age and sex standardised growth percentile* (bin width= 2 percentiles). If errors were distributed evenly throughout the distribution we would expect a uniform distribution. *Based on the modified CDC z-scores.

**Figure 4.**
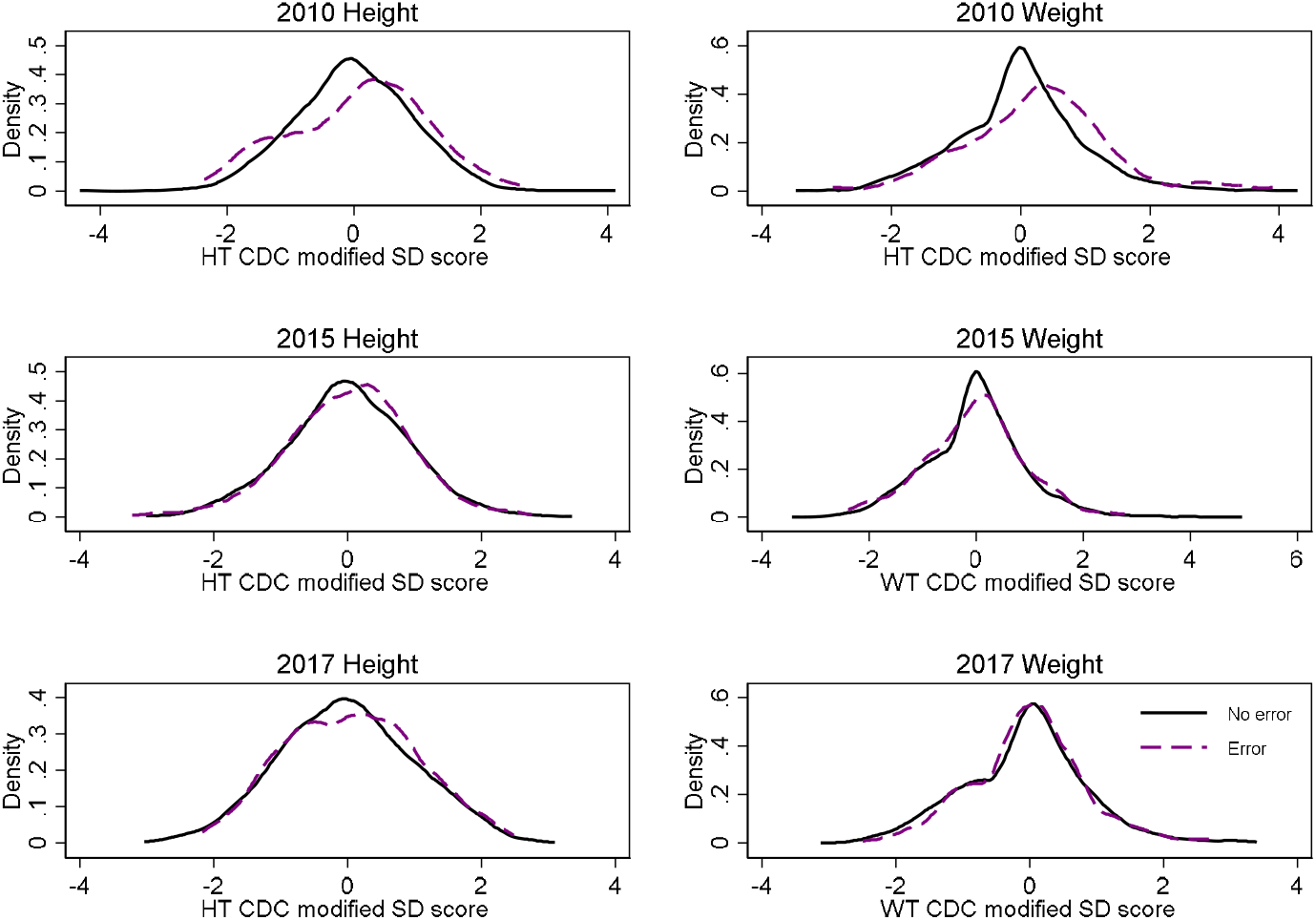
Distribution of most contemporary HT or WT z-score among children without a detected error (black line) and with an error (purple dash). The lines are Kernel density estimates.

Figure S3 (OSM file 1) shows the distribution of errors by county of residence as estimated from MLMs. There was some evidence of between county variation in cohorts 2010 and 2015 but any differences were seen in the smallest counties. With respect to the patterns of errors by maternal age, parity, education and parental birth place, out of all comparisons across all cohorts adjusting for county and number of measurements, there was only weak evidence in the 2010 cohort that errors were slightly higher in children where both parents were not born in Norway.

### Comparison with WHO cross-sectional outliers

The WHO cross-sectional cut-offs flagged <1% of values across all cohorts, in the 2010 dataset this was <0.2% of values (Table 5). The sensitivity to detect errors was poor (range 0 to 55%), and the prevalence of false positives (1-positive predictive value) particularly high for datasets 2010 and 2015 where a greater proportion of the errors were inliers. The prevalence of overweight and obesity was higher and the prevalence of grade 1 thinness lower when using the WHO cleaned dataset compared to the longitudinally cleaned data, particularly in the 2015 dataset at 8y (Figure 5)

**Table 5.** Prevalence of biologically implausible values according to the WHO-based cut-offs*, and sensitivity and specificity compared to errors found in the screening algorithm.

|  | Cohort | WHO BIV<br>n (%) | Sensitivity<br>(%) | Specificity<br>(%) | PPV (%) | NPV (%) |
| --- | --- | --- | --- | --- | --- | --- |
| Height | 2010 | 44 (0.15%) | 15.2 | 99.9 | 36.4 | 99.7 |
|  | 2015 | 188 (0.5%) | 24.6 | 99.9 | 70.2 | 98.9 |
|  | 2017 | 166 (0.9%) | 54.7 | 99.9 | 84.9 | 99.3 |
| Weight | 2010 | 6 (0.02%) | 0 | 99.98 | 0 | 99.7 |
|  | 2015 | 109 (0.3%) | 20.4 | 99.9 | 64.2 | 99.3 |
|  | 2017 | 95 (0.5%) | 32.7 | 99.96 | 92.6 | 99.01 |

\*WHO cut-offs of $>5$ or $<-5$ for weight SD score and $>5$ or $<-3$ for height score. SD scores were the modified CDC SD scores described in text and used throughout.

**Figure 5.**
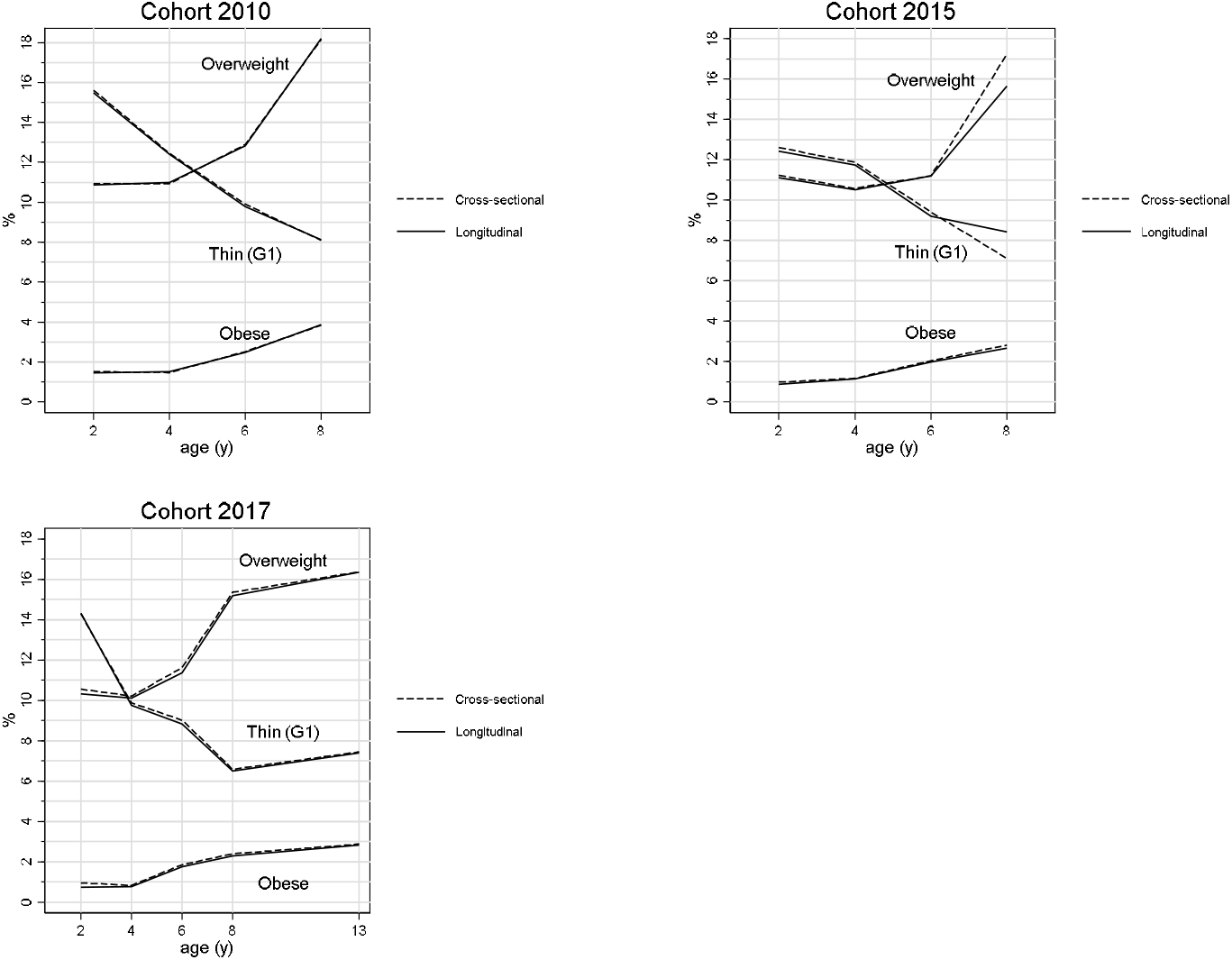
Prevalence of IOTF overweight (>25kg/m^2^), obesity (>30kg/m^2^) and grade 1 thinness (<18.5kg/m^2^) according to whether data were cleaned with the longitudinal cleaning algorithm or the cross-sectional WHO based cut-offs

## Discussion

A challenge when receiving a dataset is how to check for errors. This paper has presented a taxonomy of the different approaches reported from a scoping review, and an algorithm to systematically screen for errors in longitudinal height and weight data based on the error mechanism or way in which the error is incorrect. The latter was illustrated in three datasets where the prevalence of errors ranged from 0.44% to 2.4% of all assessments, and 4.1% to 15.3% of all children. It was not possible to automate the diagnosis of all types of error without accepting a proportion of false positives. While the screening routines were fully automated, diagnosis of errors was either not possible or over-ridden in approximately 0.5% to 3% of all children depending on the dataset. In two cohorts, the majority of errors were inliers, which meant the commonly used WHO cut-offs for error detection had poor sensitivity. Further, there were some notable differences in the prevalence of IOTF overweight and thinness when comparing the cleaning approach here with the WHO cut-off approach.

The algorithm presented here offers an automated, distribution-wide and documentable approach to screening for errors. The routines require little statistical knowledge and are perhaps more accessible compared to approaches that adopt more advanced statistical models where a mis-specified model and its consequences may be trickier for a non-statistician to diagnose. The algorithm includes checks of dates which enabled some errors to be corrected. However, these checks were rudimentary because of restricted access to the date variables. The frequency of date errors is thus likely to be substantially underestimated and many date errors were almost certainly flagged as HT or WT anomalies manifesting as unusual growth patterns.

The framework is likely to be generalisable and of use to other longitudinal datasets, particularly those which share some commonality in data pipelines, sources and structures. Some steps within the algorithm may have utility for other anthropometric measures but will require adaptation, for example, in how measurements are age- and sex-standardised.

Manual verification of the flagged values from screening and the automated diagnostic decisions was done by a statistician (author). An auxologist or endocrinologist would be preferable alongside supplemental information on the child’s medical history (17) and access to the raw data. However, definitions of what constitutes an impossible or implausible value and the principle of only removing a value if there was no uncertainty that it was an error was rigorously was adhered to. This also means that the error rates reported here should be considered as minimum estimates. Some errors almost certainly remain although these will be of smaller magnitude and of lesser consequence to validity.

### Comparison with other studies

The frequency of reported removals or flagged suspicious values in the scoping review ranged from 0.04% to ∼12.6% (3, 8, 9, 15, 17, 23, 28, 33-35, 43, 44). This is based on a multitude of different methods and thresholds. The study closest to ours in terms of using a similar multi-step algorithmic and longitudinal cleaning approach reported slightly higher error rates of ∼4 to 5% (9). The wide range of error frequencies reported most likely in large part reflect different cleaning approaches rather than true differences in data quality, as supported by this study and others that have run different routines on the same dataset (8, 9).

Few studies have reported the proportion of individuals with an error and the distribution of the number of errors per child (3, 15, 34). These statistics are useful, and the significant numbers emphasise the advantage of using multilevel growth models in substantive analyses so that allowance for measurement error is made in a principled way.

The concept of plausible outliers and implausible inliers has been outlined before (23, 45) and was reflected in the error distributions in each of our datasets. Consequently, the cross-sectional WHO cut-offs for error detection were inaccurate and caused some bias in estimates of overweight, obesity and thinness, as supported in a previous study (34). Both the sensitivity and specificity, and the bias introduced by these cross-sectional outlier approaches will depend on the error structure within the dataset, as shown in our datasets, and is likely to be difficult to predict. Distribution-wide screening with thresholds that maximise sensitivity and secondary manual validation to minimise false positives, as used here, will mitigate this. Removing true data from the tails of the distribution via selective cleaning will also cause the probability of missingness to depend on the observed value (i.e; missing not at random) (46), which may introduce bias into aetiological research.

Increasing the sophistication of the built-in rules on electronic data entry forms to include logic checks and markers of external and internal consistency could improve data quality and research efficiency by flagging and preventing errors in-situ (47). Some of the algorithm steps reported here may be applicable. Rules involving statistical models and supplementary software will be trickier to implement but as shown here and elsewhere (3) (9), there is still significant value to be gained just by implementing a set of simple cross-sectional and longitudinal logical rules.

Generally, the automated routines for diagnosing errors performed well and substantially improved the speed of data cleaning once code was produced, even with supplementary manual validation. However, it did produce some false positives, and to minimise these required manually validating 5 to 13% of the growth histories in the datasets so that incorrect decisions to be over-ruled. There is a law of diminishing returns to cleaning and this trade-off needs to be considered when designing a data cleaning strategy. The resources for developing/ implementing code, for manual review and the size of the dataset will be factors that influence the choice. Where resources are limited and the purpose is to create a shareable dataset then it may be most sensible to implement a limited number of automated checks that create no false positives.

Some of the screening steps reported here may be useful for flagging suspicious patterns for further sensitivity analyses. The types of errors that are flagged and the threshold level for flagging might depend on the research question, for example, if the interest is in change or patterns of growth, then it may be useful to run a sensitivity analysis with several different thresholds for implausible change.

These sorts of routines may also have utility in multi-cohort studies as part of a data harmonisation (5, 48) or cleaning plan as is common in genetic consortia. The metadata that flag and explain why a value was removed might also highlight differences in data quality between studies. Understanding whether errors vary by site of measurement as shown here and elsewhere (9, 32), and by other important variables may also help direct more appropriate analysis plans and aid in the interpretation of multiple cohort studies.

The characterisation of the different types of errors and their frequencies that we report might be of use to designing better simulation studies that test error detection routines. Such simulation studies should mimic the types of errors that occur in datasets, otherwise results may have little generalisability when applied to datasets that have different error structures (49). Merely simulating normally distributed errors without age (date) errors, keystroke errors (29), duplications, internally inconsistent values etc is unrealistic, and such studies are unlikely to be a useful test of the performance of the method.

There is plenty of scope to improve the algorithm and other cleaning routines more generally. Certain statistical routines may work better for screening and diagnosing particular types of errors and this is one area where performance could be optimised. Similarly integrating checks of dates with checks of size and growth will offer improvement and given the limited range of values and added complexity of dates, these are probably easier to correct and a larger source of error. Lastly, more information is needed that describes how children grow, including the distribution of normal growth and plausible growth patterns (25), this lack of reference information makes screening for implausible patterns somewhat arbitrary (3).

Work to standardise definitions, practice and reporting in cleaning could also offer benefits. Until researchers are more consistent in these practices, comparisons of error rates between studies are unlikely to accurately reflect differences in research processes and data quality or offer much insight when interpreting a substantive analysis.

Lastly, data cleaning is an arduous and time-consuming process. In some research areas it has become wrongly disincentivised under the prevailing research culture of productivity metrics and publish or perish (50-52). This runs counter to good scientific conduct and is unethical (51). Poor data cleaning, checking and quality assurance practices almost certainly contribute to the reproducibility crisis (4) and unwanted sources of between-study heterogeneity, that in the long-run are detrimental to the accrual of scientific knowledge and a waste of resources. Further, accessing information on exactly how data were collected, its quality and issues is increasingly more important as data and code are shared and made public. Its status should thus be elevated and incorporated more formally into research governance and funding, and the training of researchers (52).

In conclusion, cleaning routines for longitudinal data should incorporate longitudinal approaches to assess internal consistency and ensure distribution-wide cleaning, naive cross-sectional trimming as a stand-alone method may do more harm than good. Multi-step algorithmic approaches are recommended to systematically screen and document the wide range of ways in which errors can occur and to maximise sensitivity. Manual validation of flagged values from screening and automated diagnostic decisions are necessary to mitigate against the false removal of true values.

## Data Availability

Data are not publicly available, enquiries for access can be made to the Institute of Public Health, Norway. Statistical code is available on request.

## Acknowledgements

Ingunn Holden Bergh; Bente Øvrebø; and Ingvild Bokn provided information about the datasets used in this report. Carrie Daymont provided some updated statistical code for the SD smoothing method which was adapted for part of this work (ref #9). I am grateful to the Youth Growth research team (Elling Bere, Ingunn Holden Bergh; Per Magnus, Bente Øvrebø, Tonje Holte Stea, Pål Surén), and Rebecca Hardy and Dara O’Neill who read and provided feedback on an earlier drafts of this paper. We are grateful to the school health nurses for collection of data and all of the children and parents who participated in the Norwegian Child and Youth Growth studies.

## References

1. Non communicable Disease Risk Factor Collaboration. Height and body-mass index trajectories of school-aged children and adolescents from 1985 to 2019 in 200 countries and territories: a pooled analysis of 2181 population-based studies with 65 million participants. The Lancet. 2020;396(10261):1511–24.

2. Neta G. SJM, Rajaraman P. Quality Control and Good Epidemiological Practice. In: Ahrens W. PI, editor. Handbook of Epidemiology New York, NY: Springer; 2014.

3. Chen S, Banks WA, Sheffrin M, Bryson W, Black M, Thielke SM. Identifying and categorizing spurious weight data in electronic medical records. The American journal of clinical nutrition. 2018;107(3):420–6.

4. Munafò MR, Nosek BA, Bishop DVM, Button KS, Chambers CD, Percie du Sert N, et al. A manifesto for reproducible science. Nature Human Behaviour. 2017;1(1):0021.

5. Jaddoe VWV, Felix JF, Andersen AN, Charles MA, Chatzi L, Corpeleijn E, et al. The LifeCycle Project-EU Child Cohort Network: a federated analysis infrastructure and harmonized data of more than 250,000 children and parents. European journal of epidemiology. 2020;35(7):709–24.

6. Association AS. Ethical guidelines for statistical practice.. Alexandria (Virginia); 1999.

7. Van den Broeck J, Cunningham SA, Eeckels R, Herbst K. Data cleaning: detecting, diagnosing, and editing data abnormalities. PLoS medicine. 2005;2(10):e267.

8. Lawman HG, Ogden CL, Hassink S, Mallya G, Vander Veur S, Foster GD. Comparing Methods for Identifying Biologically Implausible Values in Height, Weight, and Body Mass Index Among Youth. American journal of epidemiology. 2015;182(4):359–65.

9. Daymont C, Ross ME, Russell Localio A, Fiks AG, Wasserman RC, Grundmeier RW. Automated identification of implausible values in growth data from pediatric electronic health records. J Am Med Inform Assoc. 2017;24(6):1080–7.

10. Munn Z, Peters MDJ, Stern C, Tufanaru C, McArthur A, Aromataris E. Systematic review or scoping review? Guidance for authors when choosing between a systematic or scoping review approach. BMC medical research methodology. 2018;18(1):143.

11. WHO. Physical Status: the use and interpretation of anthropometry. Report of a WHO Expert Committee. 1995.

12. Centers for Disease Control P. Modified z-scores in the CDC growth charts 2012; (18/12/2012).

13. Kim J, Must A, Fitzmaurice GM, Gillman MW, Chomitz V, Kramer E, et al. Incidence and remission rates of overweight among children aged 5 to 13 years in a district-wide school surveillance system. American journal of public health. 2005;95(9):1588–94.

14. Lawman HG, Mallya G, Veur SV, McCoy T, Colby L, Sanders T, et al. Trends in relative weight over 1 year in low-income urban youth. Obesity. 2015;23(2):436–42.

15. Lo JC, Maring B, Chandra M, Daniels SR, Sinaiko A, Daley MF, et al. Prevalence of obesity and extreme obesity in children aged 3-5 years. Pediatr Obes. 2014;9(3):167–75.

16. Lobstein TJ, James WP, Cole TJ. Increasing levels of excess weight among children in England. International journal of obesity and related metabolic disorders : journal of the International Association for the Study of Obesity. 2003;27(9):1136–8.

17. Smith N, Coleman KJ, Lawrence JM, Quinn VP, Getahun D, Reynolds K, et al. Body weight and height data in electronic medical records of children. International journal of pediatric obesity : IJPO : an official journal of the International Association for the Study of Obesity. 2010;5(3):237–42.

18. Sturm R. Increases in morbid obesity in the USA: 2000-2005. Public Health. 2007;121(7):492–6.

19. Conde WL, Monteiro CA. Body mass index cutoff points for evaluation of nutritional status in Brazilian children and adolescents. J Pediatr (Rio J). 2006;82(4):266–72.

20. National Health and Nutrition Examination Survey. 2001–2002 data documentation, codebook, and frequencies: body measurements 2004. Available from: http://www.cdc.gov/nchs/nhanes/nhanes2001-2002/BMX_B.htm.

21. Youth Risk Behaviour Surveillance System. 2013 YRBS data user’s guide.2012 18/12/2019. Available from: ftp://ftp.cdc.gov/pub/data/yrbs/2011/YRBS_2011_National_User_Guide.pdf.

22. Field AE, Austin SB, Taylor CB, Malspeis S, Rosner B, Rockett HR, et al. Relation between dieting and weight change among preadolescents and adolescents. Pediatrics. 2003;112(4):900–6.

23. Boone-Heinonen J, Tillotson CJ, O’Malley JP, Marino M, Andrea SB, Brickman A, et al. Not so implausible: impact of longitudinal assessment of implausible anthropometric measures on obesity prevalence and weight change in children and adolescents. Annals of epidemiology. 2019;31:69-74.e5.

24. Boswell N, Byrne R, Davies PSW. Eating behavior traits associated with demographic variables and implications for obesity outcomes in early childhood. Appetite. 2018;120:482–90.

25. Freedman DS, Lawman HG, Galuska DA, Goodman AB, Berenson GS. Tracking and Variability in Childhood Levels of BMI: The Bogalusa Heart Study. Obesity. 2018;26(7):1197–202.

26. Katzow M, Messito MJ, Mendelsohn AL, Scott MA, Gross RS. The Protective Effect of Prenatal Social Support on Infant Adiposity in the First 18 Months of Life. J Pediatr. 2019;209:77–84.

27. Morita A, Ochi M, Isumi A, Fujiwara T. Association between grandparent coresidence and weight change among first-grade Japanese children. Pediatr Obes. 2019;14(8):e12524.

28. Shi J, Korsiak J, Roth DE. New approach for the identification of implausible values and outliers in longitudinal childhood anthropometric data. Annals of epidemiology. 2018;28(3):204-11.e3.

29. Woolley CSC, Handel IG, Bronsvoort BM, Schoenebeck JJ, Clements DN. Is it time to stop sweeping data cleaning under the carpet? A novel algorithm for outlier management in growth data. PloS one. 2020;15(1):e0228154.

30. Fisher JO, Serrano EL, Foster GD, Hart CN, Davey A, Bruton YP, et al. Title: efficacy of a food parenting intervention for mothers with low income to reduce preschooler’s solid fat and added sugar intakes: a randomized controlled trial. The international journal of behavioral nutrition and physical activity. 2019;16(1):6.

31. Boswell N, Byrne R, Davies PSW. An examination of children’s eating behaviours as mediators of the relationship between parents’ feeding practices and early childhood body mass index z-scores. Obes Sci Pract. 2019;5(2):168–76.

32. Phan HTT, Borca F, Cable D, Batchelor J, Davies JH, Ennis S. Automated data cleaning of paediatric anthropometric data from longitudinal electronic health records: protocol and application to a large patient cohort. Scientific reports. 2020;10(1):10164.

33. Freedman DS, Lawman HG, Pan L, Skinner AC, Allison DB, McGuire LC, et al. The prevalence and validity of high, biologically implausible values of weight, height, and BMI among 8.8 million children. Obesity. 2016;24(5):1132–9.

34. Freedman DS, Lawman HG, Skinner AC, McGuire LC, Allison DB, Ogden CL. Validity of the WHO cutoffs for biologically implausible values of weight, height, and BMI in children and adolescents in NHANES from 1999 through 2012. The American journal of clinical nutrition. 2015;102(5):1000–6.

35. Welch C, Petersen I, Walters K, Morris RW, Nazareth I, Kalaitzaki E, et al. Two-stage method to remove population- and individual-level outliers from longitudinal data in a primary care database. Pharmacoepidemiology and drug safety. 2012;21(7):725–32.

36. Yang S, Hutcheon JA. Identifying outliers and implausible values in growth trajectory data. Annals of epidemiology. 2016;26(1):77-80 e1-2.

37. Gray CL, Robinson WR. Throwing out the baby with the bathwater?: Comparing 2 approaches to implausible values of change in body size. Epidemiology. 2014;25(4):591–4.

38. van Smeden M, Lash TL, Groenwold RHH. Reflection on modern methods: five myths about measurement error in epidemiological research. Int J Epidemiol. 2020;49(1):338–47.

39. Pohlabeln H, Reineke A, Schill W. Data Management in Epidemiology. In: Ahrens W, Pigeot I, editors. Handbook of Epidemiology. New York, NY: Springer New York; 2014. p. 979–1022.

40. Cole TJ, Lobstein T. Extended international (IOTF) body mass index cut-offs for thinness, overweight and obesity. Pediatr Obes. 2012;7(4):284–94.

41. Tanner JM, Davies PS. Clinical longitudinal standards for height and height velocity for North American children. J Pediatr. 1985;107(3):317–29.

42. Group WMGRS. WHO Child Growth Standards: Growth Velocity Based on Weight, Length and Head Circumference: Methods and Development. Geneva, Switzerland: World Health Organisation; 2009.

43. Crowe S, Seal A, Grijalva-Eternod C, Kerac M. Effect of nutrition survey ‘cleaning criteria’ on estimates of malnutrition prevalence and disease burden: secondary data analysis. PeerJ. 2014;2:e380.

44. Thurber KA, Banks E, Banwell C. Approaches to maximising the accuracy of anthropometric data on children: review and empirical evaluation using the Australian Longitudinal Study of Indigenous Children. Public Health Res Pract. 2014;25(1).

45. Daymont C. Plausible Outliers and Implausible Inliers. Obesity. 2020;28(7):1174.

46. White IR, Royston P, Wood AM. Multiple imputation using chained equations: Issues and guidance for practice. Statistics in medicine. 2011;30(4):377–99.

47. Onyango AW, Pinol AJ, de Onis M. Managing data for a multicountry longitudinal study: experience from the WHO Multicentre Growth Reference Study. Food Nutr Bull. 2004;25(1 Suppl):S46–52.

48. Johnson W, Li L, Kuh D, Hardy R. How Has the Age-Related Process of Overweight or Obesity Development Changed over Time? Co-ordinated Analyses of Individual Participant Data from Five United Kingdom Birth Cohorts. PLoS medicine. 2015;12(5):e1001828. discussion e.

49. Boulesteix AH, S; Charlton, A; Seibold, H. A replication crisis in methodological research? Significance. 2020;17(5):18–21.

50. Aitkenhead D. Peter Higgs: I wouldn’t be productive enough for today’s academic system. Guardian. 2013.

51. Frith U. Fast Lane to Slow Science. Trends Cogn Sci. 2020;24(1):1–2.

52. Mons B. Invest 5% of research funds in ensuring data are reusable. Nature. 2020;578(7796):491.

