## Supplemental File 1 for "Screening & diagnosing errors in longitudinal measures of body size"

**Online Supplementary Material File 1**

1. **Scoping Review**

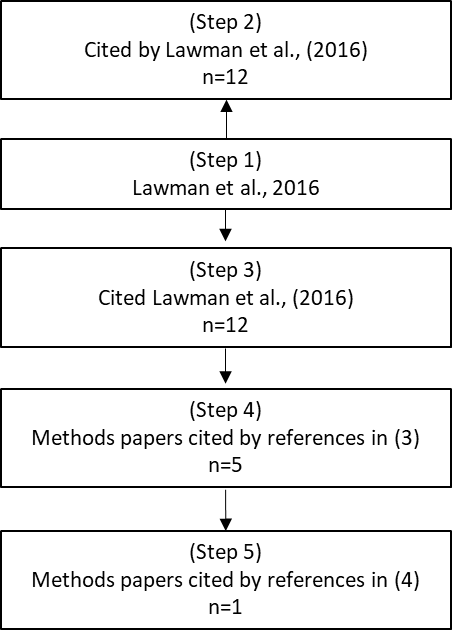

Figure S1. Schematic of literature search for scoping methods used to clean longitudinal height and weight data. Excludes citations from editorials.

References:

Step 1 (1)

Step 2 (2-13)

Step 3 (14-25)

Step 4 (26-30)

Step 5 (31)

Table S1. Taxonomy of different approaches used to clean height and weight or body mass index

| **Design of method** | **Marker/ indicator used to screen/ diagnose errors** | **Reference for threshold:** | **How was threshold selected?** | **Example(s)** |
| --- | --- | --- | --- | --- |
| Cross-sectional  trimming | Absolute value (Xi) | Biologically impossible size | Impossible limits | BMI >40 or BMI<8kg/m^2^ (6)  HT<30.5cm (7)  WT<4.5kg (5) |
|  |  | Internally derived | Arbitrary statistical-based limits | \|WTi\|> 99th percentile+200% of the median (7)  HT< 1^st^ percentile HT for age minus 30.5cm (7)  Dixon test (29); |
|  | Age & sex standardised values (SDi) | External population | Arbitrary statistical-based limits | \|Modified CDC SDi\|>WHO threshold  (2, 9, 27)\|CDC SDi\|>6 (3) |
|  |  | External population | Biologically impossible limits | \|SDi\|>25 (17) |
|  |  | Internally derived | Arbitrary statistical-based limits | \|SDi\|>3 (29) |
|  | Data entry error logic checks | Logical inconsistencies | Based on potential data entry error mechanisms | HT-WT switch; 2 values for HT entered (17); digit errors, transposed digits (22) |
| Longitudinal  trimming | Absolute change (ΔX) |  | Biologically impossible change | ΔHT<-1 inch (3) |
|  |  | External population | Arbitrary statistical-based limits | ΔX greater than value corresponding to 3 z-scores on Tanner or WHO growth velocity charts (17) |
|  | Age & sex standardised values (SDi) | External population | Arbitrary statistical-based limits | ΔHT>3SD, \|ΔBMI\|>3SD (3) |
|  |  | Internally derived | Arbitrary statistical-based limits | Top and bottom 1% of BMI change (8). |
|  | Relative change (% ΔX) | Internally derived | Based on non-linear part of the cumulative distribution of marker | % ΔX>threshold for WT gain and loss (16) |
|  | Ratio of Euclidean distances between 3 measures | Internally derived | Based on non-linear part of the cumulative distribution of marker | \|Ratio of distances\|> threshold; \|Difference in distances\|> threshold (16) |
|  | Error residuals estimated from multilevel growth model (Eij) | Internally derived | Arbitrary statistical-based limits | \|Studentised Eij\|>6 based on linear piece-wise linear spline MLM (14); \|Studentised Eij\| exceeding ordinal range of thresholds reflecting increasing likelihood of implausibility (29) |
|  | Conditional growth (Xi\|Xi-1) estimated from multilevel model | Internally derived | Arbitrary statistical-based limits | \|(Xi\|Xi-1)\|>4SD based on MLM linear spline model (30) |
|  | Jackknife residual estimated from ordinary regression on each individual | Internally derived | Arbitrary statistical-based limits | \|Jackknifed residual\| > 5 based on regression of X for each child as a function of age (21) |
|  | Deviation in SD score from individual’s smoothed (expected) SD trajectory. | Internally derived | iteratively selected conservative cut-points based on judgement of author | Deviation of CDC SD score from individual’s exponentially weighted moving average trajectory (17) |
|  | Data entry error logic checks | Logical inconsistencies | Based on potential data entry error mechanisms | Measures carried forward (17) |

CDC: Centre for Disease Control and Prevention; MLM: Multi-level model;

2. **Data harmonisation**

Information at birth was available from both the Medical Birth Registry (MBR) and Health cards. There was only small disagreement between sources, for example, <1% of observations disagreed by more than 200g for birth weight and >1cm for birth length. However, the MBR data contained more children and is the more widely used and official record of birthweight, and so was kept and health card information used only when there was no MBR record.

The clothing correction was applied to the school clinic data to align with the health card data as per the National Measurement guidelines. This produces the greatest degree of harmonisation between and within cohorts.

Information at 8yrs of age was available from both the school research clinic and health card in the 2010 (n=52) and 2015 cohorts (n=1158). To harmonise the structure of primary and secondary measurements around this age between cohorts with respect to the different information sources, measurements from the health card were removed if they occurred within 9 months of the school clinic.

**3 Data Structure**

Table S2 Number of observations by measurement schedule

|  | **2010** |  | **2015** |  | **2017** |  |
| --- | --- | --- | --- | --- | --- | --- |
| **Target age** | **n** | **%** | **n** | **%** | **n** | **%** |
| birth | 3182 | 10.2 | 3076 | 8.1 | 1741 | 9.4 |
| 1-5d (2-4d) | 7 | 0.0 | 20 | 0.1 | 13 | 0.1 |
| 6-21d (7-10d) | 25 | 0.1 | 170 | 0.4 | 22 | 0.1 |
| 6wks (21-67d) | 2525 | 8.1 | 2607 | 6.8 | 1585 | 8.6 |
| 3m (68-137d) | 2655 | 8.5 | 4321 | 11.3 | 1697 | 9.2 |
| 5m (138-167d) | 308 | 1.0 | 2078 | 5.4 | 1120 | 6.1 |
| 6m (168-243d) | 2657 | 8.6 | 3888 | 10.2 | 464 | 2.5 |
| 10m (244-335d) | 2484 | 8.0 | 3379 | 8.8 | 52 | 0.3 |
| 12m (336-446d) | 2791 | 9.0 | 3044 | 8.0 | 1665 | 9.0 |
| 15-18m (447-640d) | 3267 | 10.5 | 3039 | 8.0 | 1572 | 8.5 |
| 2y (641-914d) | 2444 | 7.9 | 2268 | 5.9 | 1374 | 7.4 |
| 3y (915-1278d) | 618 | 2.0 | 669 | 1.8 | 257 | 1.4 |
| 4y (1279-1643d) | 2102 | 6.8 | 2570 | 6.7 | 1490 | 8.1 |
| 5/6y (1644-2557d) | 2768 | 8.9 | 3420 | 9.0 | 1794 | 9.7 |
| 8y (2558-3653d) Health Card | 52 | 0.2 | 299 | 0.8 | 1678 | 9.1 |
| 8y (2558-3653d) School | 3182 | 10.2 | 3335 | 8.7 | 0 | 0.0 |
| 13y (3654-5844d) School | 0 | 0.0 | 0.0 | 0.0 | 1892 | 10.2 |

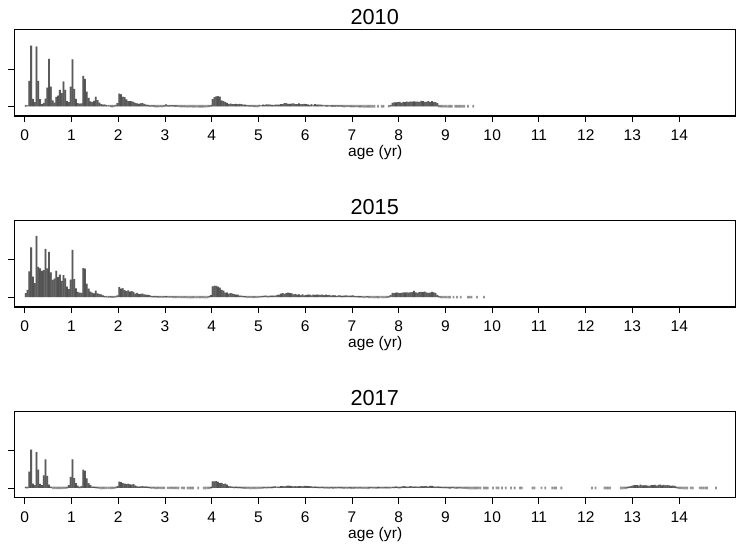

Figure S2. Histograms of measurement ages in each cohort

Table S3. Frequency of number of measures per child in each cohort.

| N measures | Cohort 2010 | | | | Cohort 2015 | | | | Cohort 2017 | | | |
| --- | --- | --- | --- | --- | --- | --- | --- | --- | --- | --- | --- | --- |
|  | Weight |  | Height |  | Weight |  | Height |  | Weight |  | Height |  |
|  | n | Cum. % | n | Cum. % | n | Cum. % | n | Cum. % | n | Cum. % | n | Cum. % |
| 0 | 0 | 0 | 0 | 0 | 0 | 0 | 0 | 0 | 1 | 0,1 | 0 | 0,0 |
| 1 | 116 | 3,6 | 122 | 3,8 | 56 | 1,7 | 64 | 1,9 | 45 | 2,4 | 45 | 2,4 |
| 2 | 130 | 7,7 | 124 | 7,7 | 216 | 8,1 | 208 | 8,1 | 63 | 5,7 | 63 | 5,7 |
| 3 | 5 | 7,9 | 5 | 7,9 | 104 | 11,3 | 107 | 11,4 | 25 | 7,0 | 27 | 7,1 |
| 4 | 11 | 8,2 | 17 | 8,4 | 129 | 15,1 | 128 | 15,2 | 24 | 8,3 | 23 | 8,3 |
| 5 | 36 | 9,4 | 43 | 9,8 | 108 | 18,4 | 106 | 18,4 | 27 | 9,7 | 27 | 9,7 |
| 6 | 64 | 11,4 | 74 | 12,1 | 76 | 20,6 | 81 | 20,8 | 20 | 10,7 | 24 | 11,0 |
| 7 | 76 | 13,8 | 102 | 15,3 | 44 | 22,0 | 45 | 22,1 | 33 | 12,5 | 40 | 13,1 |
| 8 | 124 | 17,7 | 188 | 21,2 | 65 | 23,9 | 60 | 23,9 | 48 | 15,0 | 86 | 17,6 |
| 9 | 295 | 26,9 | 473 | 36,1 | 58 | 25,6 | 60 | 25,7 | 148 | 22,8 | 208 | 28,5 |
| 10 | 720 | 49,5 | 772 | 60,3 | 116 | 29,1 | 124 | 29,4 | 461 | 46,9 | 523 | 55,9 |
| 11 | 1009 | 81,2 | 877 | 87,9 | 186 | 34,7 | 186 | 35,0 | 987 | 98,7 | 822 | 99,0 |
| 12 | 495 | 96,8 | 327 | 98,1 | 348 | 45,1 | 348 | 45,4 | 25 | 100,0 | 19 | 100,0 |
| 13 | 99 | 99,9 | 59 | 100,0 | 431 | 58,0 | 433 | 58,4 | 0 | 100,0 | 0 | 100,0 |
| 14 | 3 | 100,0 | 0 | 100,0 | 450 | 71,5 | 448 | 71,8 | 0 | 100,0 | 0 | 100,0 |
| >14 | 0 | 100,0 | 0 | 100,0 | 951 | 100,0 | 940 | 100,0 | 0 | 100,0 | 0 | 100,0 |
| Total | 3183 | 100 | 3183 | 100 | 3338 | 100 | 3338 | 100 | 1907 | 100 | 1907 | 100 |

Table S4. Glossary of terms for describing cleaning processes (adapted from Van den Broeck et al., 2004(28))

| Data flow | Passage of data from measurement to analysis dataset |
| --- | --- |
| Quality control | Activities that occur during or after data collection to correct data errors |
| Data cleaning | Dealing with problems once they have occurred in the database |
| Inlier | A data value within an acceptable range |
| Outlier | A data value considered extreme |
| Impossible value | A data value that cannot be true |
| Implausible value | A data value that is subjectively extremely unlikely |
| Suspicious value | A data value that is possible but has a very low probability of occurring. |
| Analysis dataset | Dataset after cleaning |

Table S5. Frequency of duplicates in each cohort

|  | **2010** | **2015** | **2017** |
| --- | --- | --- | --- |
| N (%) | 117 (0.4%) | 772 (2.0%) | 86 (0.5%) |
| Total Assessments | 31067 | 38183 | 18474 |

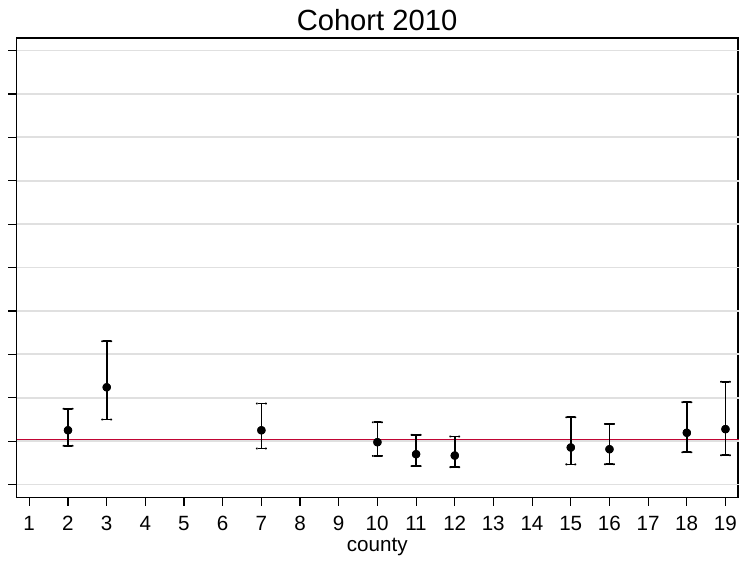

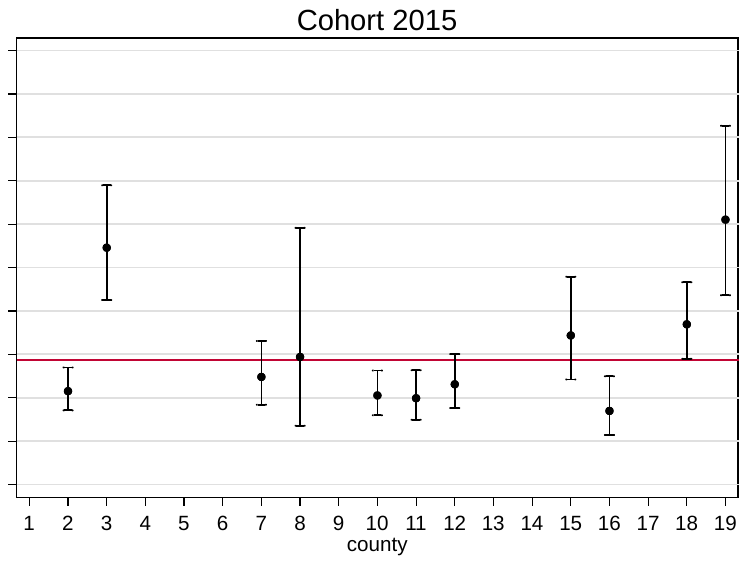

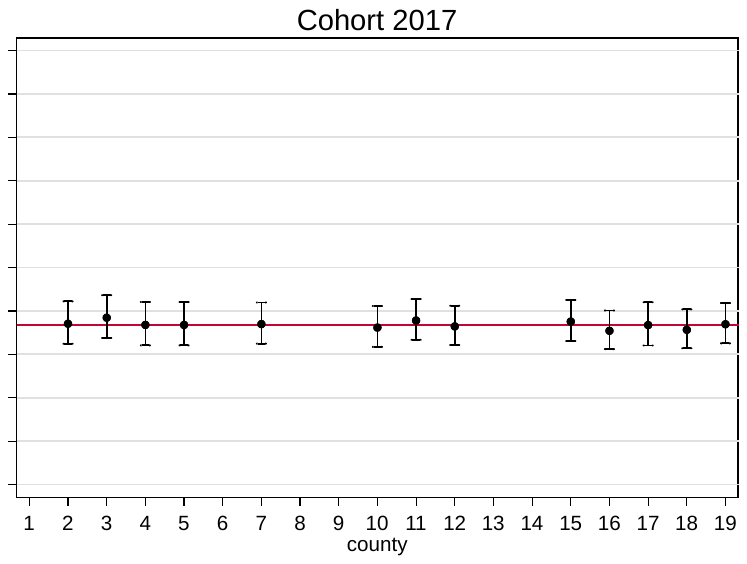

Figure S3. Predicted % of children with errors by county as estimated from multilevel logistic models (adjusted for number of measurements).

References

1.	Lawman HG, Ogden CL, Hassink S, Mallya G, Vander Veur S, Foster GD. Comparing Methods for Identifying Biologically Implausible Values in Height, Weight, and Body Mass Index Among Youth. American journal of epidemiology. 2015;182(4):359-65.

2.	Centers for Disease Control P. Modified z-scores in the CDC growth charts2012; (18/12/2012).

3.	Kim J, Must A, Fitzmaurice GM, Gillman MW, Chomitz V, Kramer E, et al. Incidence and remission rates of overweight among children aged 5 to 13 years in a district-wide school surveillance system. American journal of public health. 2005;95(9):1588-94.

4.	Lawman HG, Mallya G, Veur SV, McCoy T, Colby L, Sanders T, et al. Trends in relative weight over 1 year in low-income urban youth. Obesity. 2015;23(2):436-42.

5.	Lo JC, Maring B, Chandra M, Daniels SR, Sinaiko A, Daley MF, et al. Prevalence of obesity and extreme obesity in children aged 3-5 years. Pediatr Obes. 2014;9(3):167-75.

6.	Lobstein TJ, James WP, Cole TJ. Increasing levels of excess weight among children in England. International journal of obesity and related metabolic disorders : journal of the International Association for the Study of Obesity. 2003;27(9):1136-8.

7.	Smith N, Coleman KJ, Lawrence JM, Quinn VP, Getahun D, Reynolds K, et al. Body weight and height data in electronic medical records of children. International journal of pediatric obesity : IJPO : an official journal of the International Association for the Study of Obesity. 2010;5(3):237-42.

8.	Sturm R. Increases in morbid obesity in the USA: 2000-2005. Public Health. 2007;121(7):492-6.

9.	WHO. Physical Status: the use and interpretation of anthropometry. Report of a WHO Expert Committee. 1995.

10.	Conde WL, Monteiro CA. Body mass index cutoff points for evaluation of nutritional status in Brazilian children and adolescents. J Pediatr (Rio J). 2006;82(4):266-72.

11.	National Health and Nutrition Examination Survey. 2001–2002 data documentation, codebook, and frequencies: body measurements2004. Available from: <http://www.cdc.gov/nchs/nhanes/nhanes2001-2002/BMX_B.htm>.

12.	Youth Risk Behaviour Surveillance System. 2013 YRBS data user’s guide.2012 18/12/2019. Available from: <ftp://ftp.cdc.gov/pub/data/yrbs/2011/YRBS_2011_National_User_Guide.pdf>.
