## Supplemental File 2 for "Screening & diagnosing errors in longitudinal measures of body size"

**ONLINE SUPPLEMENTARY FILE 2**

**OUTLINE OF SCREENING ALGORITHM STEPS**

**PART A DATES**

*Target:	Impossible dates within the context of the study and participant time-frame*

(1) Is date RECOGNISABLE?
(a) Does day, month & year vars ALL have plausible values?
(b) Is there a missing value for day or month or year?
(2) Is year within the STUDY TIME-FRAME?
(a) Does millennium take the value “2”?
(b) Does CENTURY take the value 0?
(c) Does decade or year take a value such that it falls within STUDY TIME-FRAME?
(3) Is year within PARTICIPANT TIME-FRAME?
(a) Are three or more ages<0?

Mechanism: measurements assigned to the wrong child (footnote 1)

(b) Has birth year been recorded instead of year of assessment?
(c) Is date still before PARTICIPANT TIME-FRAME?
(d) Is date after PARTICIPANT TIME-FRAME? (footnote 1)

**PART B IDENTIFIABLE DATA ENTRY ERRORS**

*Target:	Errors that can be linked to a data entry error mechanism.*

(1) DUPLICATE BIRTH MEASUREMENTS (footnote 1)

(a) DUPLICATE BIRTH LENGTH AND BIRTH WEIGHT but NOT AGE?

(b) DUPLICATE BIRTH WEIGHT ONLY?
(c) DUPLICATE BIRTH LENGTH ONLY?
(2) HT and WT values SWITCHED?
(3) HT or WT ENTERED TWICE?
(4) INCORRECT MEASUREMENT UNITS?
(5) DUPLICATE with a value outside the age-related size threshold (footnotes 2, 3)
(a) DUPLICATE HT and WT outside age-related threshold?
(b) DUPLICATE WT outside age-related threshold?
(c) DUPLICATE HT outside age-related threshold?
(d) DUPLICATE AGE (assessment date) & HT OR WT outside age-related threshold?
(6) Values SHRUNK by a factor of 10?

Mechanism:	Decimal point entered in wrong place or a trailing zero not entered (eg; 1.8kg instead of 18kg for WT, 5.5cm v 55cn for HT)

Approach:	Values indicating this error mechanism were corrected and checked for INTERNAL CONSISTENCY with other values from the individual’s growth set (footnote 4a). If they were not INTERNALLY CONSISTENT, they were removed.

(7) Values INFLATED by a factor of 10?

Mechanism:	A decimal point was omitted (eg; 65kg instead of 6.5kg for infant WT, 705cm instead of 70.5cm for HT).

Approach:	Values indicating this error mechanism were corrected and checked for INTERNAL CONSISTENCY with other values from the individual’s growth set (footnote 4a). If they were not INTERNALLY CONSISTENT, they were removed.

(8) WT values INFLATED by a factor of 100? (footnote 1)

Mechanism:	A decimal point was omitted (eg; 1023kg entered instead of 10.23kg for WT)

Approach:	Values indicating this error mechanism were corrected and checked for INTERNAL CONSISTENCY with other values from the individual’s growth set (footnote 4a). If they were not INTERNALLY CONSISTENT, they were removed.

(9) IMPOSSIBLE ABSOLUTE AGE-RELATED VALUES:

Approach:	Remove IMPOSSIBLE HT or WT values that are outside of the plausible age-related thresholds and cannot be corrected (footnotes 2,3) (nb; measures in first year of early preterms <32weeks not screened as we can expect some very low values)

(10) DUPLICATE HT & WT values where both HT & WT are INTERNALLY INCONSISTENT?

Approach:	Remaining duplicate HT & WTs are screened and a threshold is set for diagnosing the duplicate pair as INTERNALLY INCONSISTENT based on the difference between the duplicate value and the mean modified SD score for the individual (footnote 4a)

(11) HALF RECORD DUPLICATES ON THE SAME DATE

Approach:	Screen duplicate values that share the same assessment date. Jackknife residuals (footnote 4b) used to diagnose the erroneous duplicate.

(a) DUPLICATE AGE and WT
(b) DUPLICATE AGE and HT
(c) DUPLICATE AGE ONLY
(12) DUPLICATE HT AND/OR WT BUT ON DIFFERENT DATES

Approach:	Screen remaining DUPLICATES not dealt with in B1 (duplicates involving birth), B5 (impossible duplicates), B10 (internally inconsistent HT & WT duplicates) or B11 (duplicates involving age)

(a) DUPLICATE HT and WT

Approach:	Duplicate HT and WT pairs on different dates are not biologically impossible but are implausible. Jackknife residuals (footnote 4b) were used to diagnose the most internally inconsistent duplicate.

(b) DUPLICATE WT only:

Approach:	Duplicate WTs on their own are NOT biologically impossible but are implausible if they occur over a long time period and/or alongside concomitant increases in HT. The aberration to the growth velocity of the individual before and after the duplicate pair was used to diagnosis the most internally inconsistent (erroneous) duplicate (footnote 4c)

(c) DUPLICATE HT only:

Approach:	Duplicate HTs on their own are NOT biologically impossible but are implausible if they occur over a long time period and/or alongside concomitant increases in WT. The aberration to the growth velocity of the individual before and after the duplicate pair was used to diagnosis the most internally inconsistent (erroneous) duplicate (footnote 4c)

**C BIOLOGICALLY IMPOSSIBLE/ IMPLAUSIBLE CHANGE**

*Target:	Values that exceed thresholds for impossible or implausible absolute and relative change (patterns of growth)*

(1) Multiple IMPOSSIBLE HT DECREASES

Mechanism(s):	Incorrect date OR ID incorrectly assigned.

Approach:	These mainly manifest as spikes in the growth curve & are often accompanied by similar patterns in WT. A combination of visual inspection of plots and internal consistency of modified CDC z-scores (footnote 4a) was used to detect the errors among the flagged decreases. Where there were ≥3 HT decreases, it was impossible to diagnose the errors and so all values for that individual are diagnosed as errors. This may have occurred if the values were assigned to the wrong ID (child).

(2) IMPOSSIBLE HT DECREASES

Mechanism(s):	Date or data entry error.

Approach:	These mainly manifest as spikes in the growth curve & are often accompanied by similar patterns in WT suggesting a date error. Jackknife residuals were used to identify and remove the erroneous value (footnote 4b). Both HT & WT were removed if a HT decrease was accompanied by WT decrease. The routine was looped because sometimes two errors followed each other.

(3) IMPOSSIBLE HT INCREASES
(a) DUPLICATE with IMPOSSIBLE HT INCREASE

Mechanism:	Date or data entry error.

Approach:	This step screens duplicates that are <90 days apart (ie, those not screened in step B12). Duplicate values that cause an increase in HT exceeding a threshold for impossible absolute HT growth (footnote 3b) are flagged. Modified SD scores were used to quantify the internal consistency of the flagged values and to diagnose the erroneous value causing the impossible increase (footnote 4a).

(b) IMPOSSIBLE HT & WT INCREASE

Mechanism:	Date, data entry error or ID incorrectly assigned.

Approach:	Values exceeding a threshold for impossible absolute HT increase (footnote 3b) & exceeding a threshold for change in WT SD (footnote 3x) were flagged (nb; flagged values from preterm babies in the first year of life are not included to avoid conflating error with expected catchup growth). Modified SD scores were used to quantify the internal consistency of the flagged values and to diagnose the erroneous value causing the impossible increase (footnote 4a).

(c) IMPOSSIBLE HT INCREASE ONLY

Mechanism:	Date or data entry error.

Approach:	Values exceeding a threshold for impossible absolute HT increase (footnote 3b) were flagged (nb; flagged values from preterm babies in the first year of life are not included to avoid conflating error with expected catchup growth). Modified SD scores were used to quantify the internal consistency of the flagged values and to diagnose the erroneous value causing the impossible increase (footnote 4a).

(4) IMPOSSIBLE/ IMPLAUSIBLE HT (& WT) PATTERNS (footnote 1)
(a) IMPOSSIBLE/ IMPLAUSIBLE HT & WT PATTERNS

Mechanism:	Date, data entry error or ID incorrectly assigned.

Approach:	Values for both ΔHT SD & ΔWT SD that exceed thresholds for ΔSD (footnote 3c) were flagged (nb; flagged values from preterm babies in the first year of life are not included to avoid conflating error with expected catchup growth). Modified SD scores were used to quantify the internal consistency of the flagged values and to diagnose the erroneous value causing the impossible/implausible pattern (footnote 4a).

(b) IMPOSSIBLE HT PATTERN ONLY

Mechanism:	Date or data entry error.

Approach:	Values for ΔHT SD that exceed thresholds for ΔSD (footnote 3c) were flagged (nb; flagged values from preterm babies in the first year of life are not included to avoid conflating error with expected catchup growth). Modified SD scores were used to quantify the internal consistency of the flagged values and to diagnose the erroneous value causing the impossible/implausible pattern (footnote 4a).

(5) IMPOSSIBLE WT PATTERN DECREASE

Mechanism:	Date or data entry error.

Approach:	Values for ΔWT SD decreases below a threshold for ΔSD (footnote 3c) are flagged (nb; patterns of immediate postnatal weight loss are expected and ignored). Modified SD scores were used to quantify the internal consistency of the flagged values and to diagnose the erroneous value causing the impossible/implausible pattern (footnote 4a).

(6) IMPOSSIBLE WT PATTERN INCREASE

Approach:	Values for ΔWT SD increases exceeding a threshold for ΔSD (footnote 3c) are flagged (nb; flagged values from preterm babies in the first year of life are not included to avoid conflating error with expected catchup growth). Modified SD scores were used to quantify the internal consistency of the flagged values and to diagnose the erroneous value causing the impossible/implausible pattern (footnote 4a).

**FOOTNOTES**

1. Additional information for particular steps:

| **STEP** | **DETAILS** |
| --- | --- |
| A3(a) | ALL observations for child removed as measurements were suspected to have been assigned to wrong child. |
| A3(d) | This correction could be wrong. However, it is assessed for internal consistency using HT & WT in Part C of the algorithm, and if inconsistent the corrected value is removed. |
| B1 | Duplicate values from birth spaced >5weeks apart were assumed to be errors where the birth measure(s) had been carried forward or incorrectly entered twice. |
| B8 | A check of whether HT (cm) values were INFLATED by a factor of 100 could also be included here. However, such errors require both an extra digit AND a missing decimal place. Checks in our data showed that this did not occur. |
| C1 | When there are multiple errors per individual as may be caused by an ID switch, the error load is high (see footnote 2 below) and automated detection of incorrect values based on internal consistency performs poorly. However, the number of individuals with these types of multiple errors was small and so it is manageable to manually clean using visual and statistical guides to aid the decision. |
| C4 | These did not manifest as spikes as spikes are picked up in C1-3 but show as periods of impossible/ implausible change over longer periods. |

2. High error load

The validity of automated longitudinal error detection routines that are based on comparing values with summary statistic(s) derived from the same individual (within-individual) (eg; see routines (a) to (d) in footnote 4) is very sensitive to the number and magnitude of errors (error load) within an individual.

It was thus necessary to reduce the error load before executing any automated longitudinal statistical routines on flagged values such as the Jackknife residual (see footnote 3). For example, parts B5 and B9 of the algorithm which remove impossible values based on absolute thresholds were predominately positioned to reduce the error load before the longitudinal routines in steps B11 & B12

3. Notes on thresholds
(a) Thresholds for absolute impossible values:

The age-related thresholds to define an impossible value are given below (Table 1). International growth curves were used to initially select the thresholds, these were then adjusted iteratively by visualising the growth histories and checking the data of flagged individuals to ensure the thresholds did not remove true values.

Table 1. Age-related size thresholds used to define plausible values for algorithm.

|  |  | Weight |  |  |  |  | Height |  |  |
| --- | --- | --- | --- | --- | --- | --- | --- | --- | --- |
|  | Age Range (days) | | Thresholds (kg) | |  | Age Range (days) | | Thresholds (cm) | |
| Age Interval | Min | Max | lower | upper |  | Min | Max | lower | upper |
| Birth to 10 days | 0 | 10 | 0.61 | 6.7 |  | 0 | 10 | 30 | 61 |
| 11 days to 6m | 11 | 183 | 1.2 | 12 |  | 11 | 183 | 40 | 79 |
| 6m to 2y | 183 | 731 | 4 | 19 |  | 183 | 731 | 55 | 100 |
| 2y to 5y | 731 | 1826 | 7.5 | 32 |  | 731 | 1826 | 70 | 135 |
| 5 to 9y | 1826 | 3287 | 9.8 | 62 |  | 1826 | 3287 | 88 | 160 |
| 9y to 14y | 3287 | 5114 | 12.3 | 120 |  | 3287 | 5114 | 100 | 200 |

(b) Thresholds for absolute impossible HT increase

Age-related thresholds to define impossible increases between two consecutive measures of HT were based on the WHO and Tanner growth velocity charts (1, 2). The thresholds were chosen to be conservatively high through an iterative process to ensure we flagged all impossible increases but minimised false positives which were over-ruled with secondary manual validation of growth histories. The thresholds were weighted for each individual by the age period(s) over which each pair of measures occurred. This approach, based on absolute values, was not applied to WT because WT shows more biological variability which makes it difficult to define impossible levels of absolute change over long periods of growth.

(c) Thresholds for impossible/ implausible patterns of (age and sex standardised) growth.

The modified SD scale was used to flag implausible patterns of growth that may be caused by errors. This is based on the modified SD scale as changes on the absolute scale depend on age and t and which would make defining patterns on this scale to complex, particularly for unbalanced data. Thresholds were selected iteratively by inspection of plots and ΔSD values to capture all errors manifesting as impossible/ implausible growth (100% sensitivity) but minimising the number of false positives. Larger thresholds were required for longer time intervals since larger centile crossing is more plausible over longer growth intervals (3), for example, the thresholds for 2015 HT (part C3a) were: Δt≤15 days: |ΔSD|>1.5; 14<Δt≤31days: |ΔSD|>2.0; 31days<Δt≤3m: |ΔSD|>2.5; Δt>3m: |ΔSD|>3.5.

4. Routines for checking consistency of corrected values and diagnosing errors

The following table briefly outlines the routines that were used to check the internal consistency or inconsistency of a value, or to diagnose an error from a set of flagged values. Note that ALL automated diagnoses of errors using the routines below was validated using visual inspections of growth histories and over-ridden where considered incorrect.

| (a) | **Check of the Internal consistency/ inconsistency of SD values:**  Here, consistency is quantified based on the difference between the flagged value’s modified SD-score and the mean of other non-flagged values in the individual’s dataset.  For steps B6 to B8, the purpose was to check the consistency of the “corrected value”. To do this, a consistency threshold was set iteratively using visualisations of the individual growth plots of children with the corrected value and a threshold was chosen such that no erroneous corrected values were accepted. For HT, corrected values also had to meet the criteria of not violating a pattern of monotonically increasing growth. If the corrected value was not accepted, then it was not removed but picked up as an impossible value (error) in a later step (step B9) or as impossible change in part C.  For step B10, the purpose was to diagnose errors based on the internal inconsistency of flagged HT and WT duplicates. A threshold was set based on visualisations of growth curves containing the flagged duplicates such that no true values were removed.  For step C3, the purpose was to select which value was the error from pairs of values that had been flagged as causing impossible absolute HT velocities. |
| --- | --- |
| (b) | **Jackknife residuals (4)**  Regression models that allow for non-linearity were fitted to flagged individuals, and the value with the highest Jackknife residual from the set of flagged values was diagnosed as the error (least consistent).  This routine cannot be used for flagged individuals with ≤3 observations. It also performed poorly when the error load was high, for example, in B11 & B12 for individuals with >1 duplicate or >1 pair of duplicates. In these settings the diagnosis of errors was made manually using visualisations (see above). |
| (c) | **Velocity (growth) aberration**  The value, from a set of 2 duplicates, which caused the highest change in velocity (acceleration) between successive measurements before and after the duplicate pair is diagnosed as the error. A threshold was also set to avoid removing plausible duplicates that occur over a period of slow growth.  This automated diagnostic routine could not be used when (i) duplicates were not adjacent to each another & (ii) when a duplicate was the first or last measure of an individual’s dataset and so either had no earlier or later measure from which to calculate change in velocity or growth. In these settings the erroneous duplicate(s) were diagnosed manually using visualisations. |
| (e) | **Manual Routines**  Manual screening of growth histories and statistics that are a marker of the internal consistency of a value with respect to other measures from the same individual were performed to validate all automated diagnostic decisions and minimise falsely assigning a true value as an error. Manual screening was also done for certain scenarios that were difficult to automate (described above). |

References

1.	WHO Child Growth Standards: Growth Velocity Based on Weight, Length and Head Circumference: Methods and Development. Geneva, Switzerland: World Health Organisation; 2009.

2.	Tanner JM, Davies PS. Clinical longitudinal standards for height and height velocity for North American children. J Pediatr. 1985;107(3):317-29.

3.	Freedman DS, Lawman HG, Galuska DA, Goodman AB, Berenson GS. Tracking and Variability in Childhood Levels of BMI: The Bogalusa Heart Study. Obesity. 2018;26(7):1197-202.

4.	Shi J, Korsiak J, Roth DE. New approach for the identification of implausible values and outliers in longitudinal childhood anthropometric data. Annals of epidemiology. 2018;28(3):204-11.e3.
