## Supplemental File 3 for "Screening & diagnosing errors in longitudinal measures of body size"

**Online supplementary Material File 3**

**Exemplars from screening algorithm**

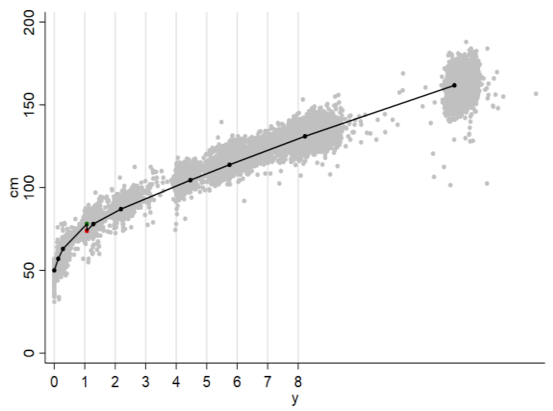


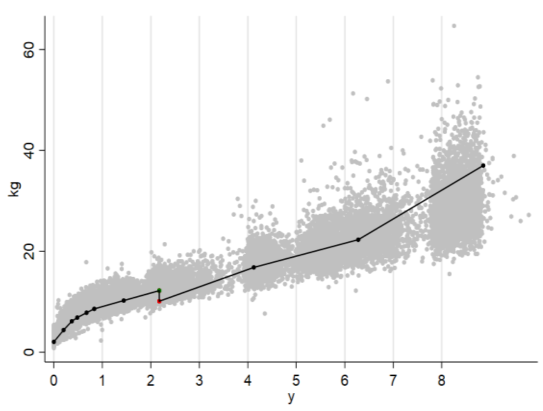


Figure S1. Exemplar plots of children with a duplicate value for AGE and WT (BUT NOT HT) (step B11a) and AGE and HT (BUT NOT WT) (step B11b). The scatter are the individual data points in the sample and are underlaid as a guide to aid manual verification.

RIGHT PLOT: The HT history of a child with a duplicate age and WT value at approximately 1-yr of age. The red marker has the highest jack-knife residual and is selected for automated removal but in this example it was over-ridden and the green value removed along with its corresponding AGE & WT duplicate.

LEFT PLOT: The WT history of a child with a duplicate age & HT value. The red marker has the highest jack-knife residual and was removed along with its corresponding AGE & HT duplicate.

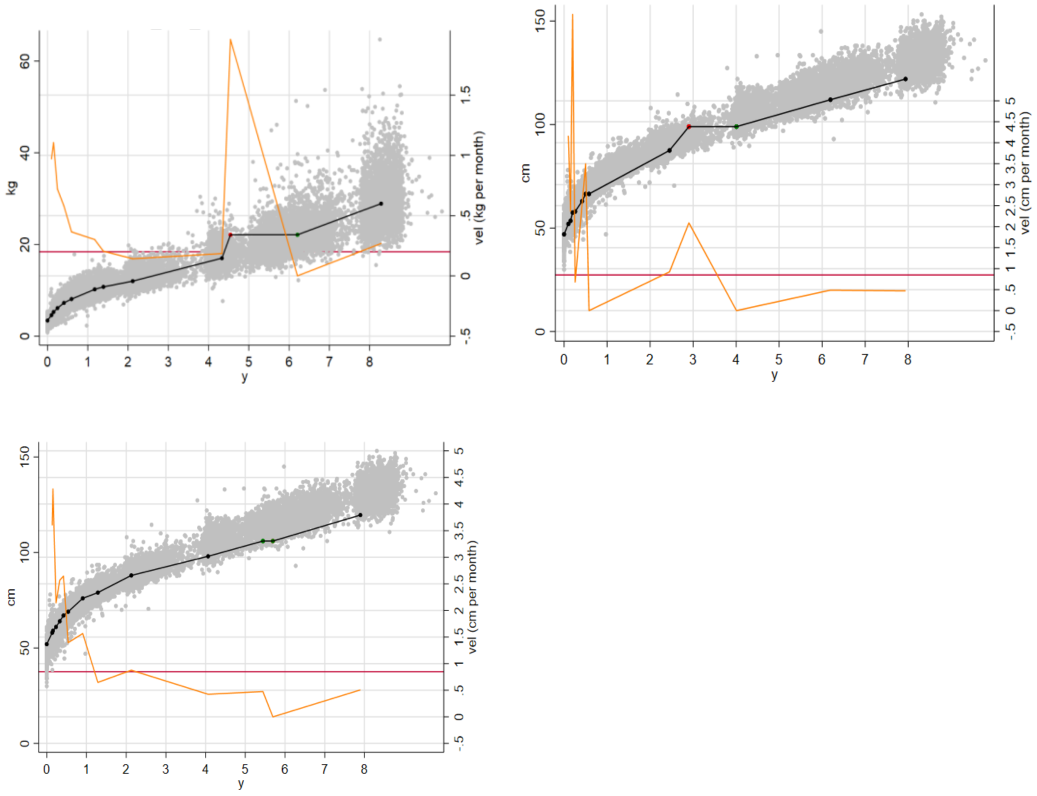


Figure S2: Exemplar plots of children that had a duplicate HT or WT value recorded on separate occasions (step B12 of algorithm). Each plot shows size (black line) and velocity (orange line) from three individuals with duplicate values on separate occasions. The red line is the threshold for velocity aberration. The scatter are the individual data points in the sample and are underlaid as a guide to aid manual verification.

TOP LEFT PLOT: Child with a duplicate WT measure, the automated routine selects the erroneous duplicate as that which causes the greatest aberration to growth velocity as the error (red marker).

TOP RIGHT PLOT: A similar example but for a child with a duplicate HT value.

BOTTOM PLOT: An example of an individual with a duplicate HT value between 5 and 6y but where the velocity aberration was below the threshold (red line) and so both duplicates are kept.

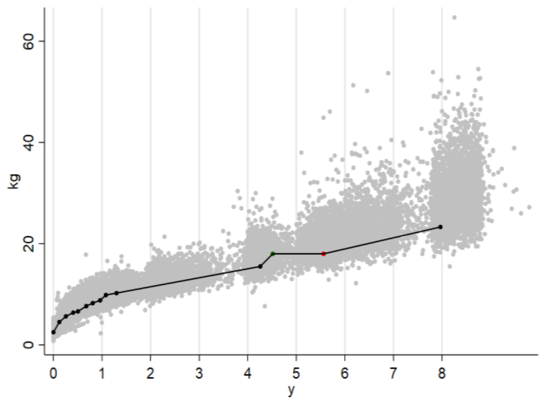


Figure S3 An exemplar of a child with a duplicate HT & WT value (step B12a) where the automated jackknife based rule selected the incorrect duplicate as the error (red dot) and the decision was over-ridden. The scatter are the individual data points in the sample and are underlaid as a guide to aid manual verification.

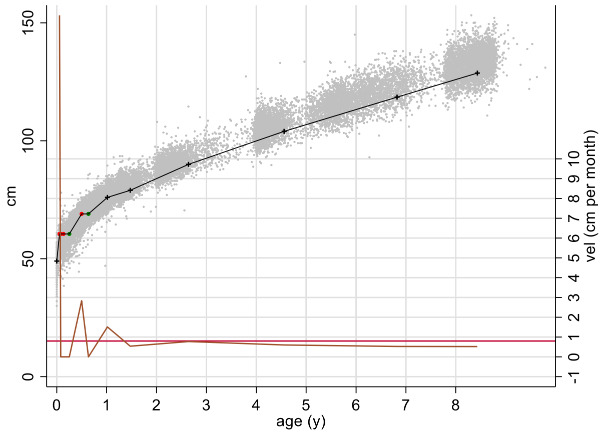


Figure S4. An exemplar plot of a child with duplicate HT values where the routine was not automated (step B12c). The plot shows size (black line) and velocity (orange line). The red line is the threshold for velocity aberration. The scatter are the individual data points in the sample and are underlaid as a guide to aid manual verification.

This child’s measurements contain two sets of duplicates for HT and the first set contains >2 duplicates, the manually diagnosed errors are coloured red and the kept observations green. Values from the rest of sample were underlaid to help verification.

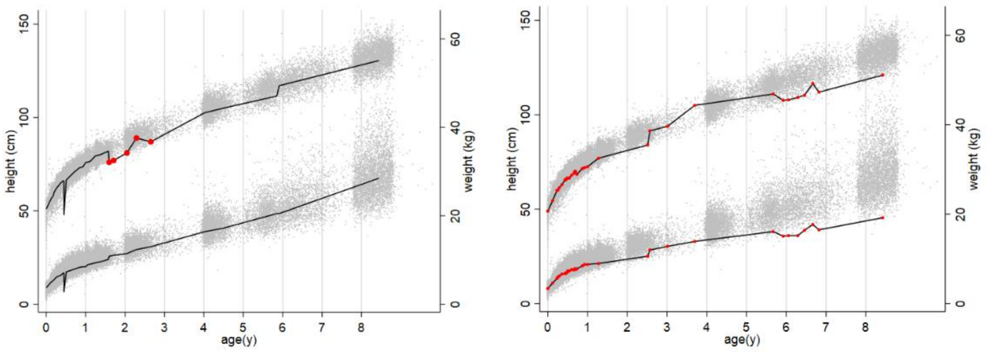


Figure S5. Exemplar plots of children with data that contained >1HT decrease (part C1). In these examples diagnosis could not be automated and so the errors were diagnosed manually. The scatter are the individual data points in the sample and are underlaid as a guide to aid manual verification.

LEFT PLOT: the measurements in red were removed. Two errors clearly remain- one co-temporal decrease with WT at approx. 6m of age and one rapid HT increase at ~6y of age. These are both flagged and addressed in part C2 and C3 of the algorithm.

RIGHT PLOT: the measurements assigned to this ID appear to come from 2 different children. Since it was not possible to tell which is correct, all values were removed.

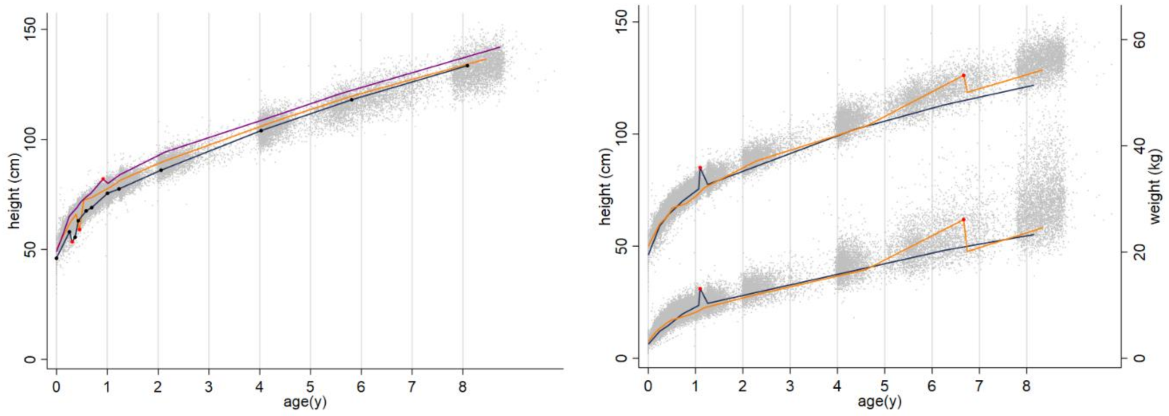


Figure S6. Exemplars of children with data that contained a single HT decrease (part C2a&b). The jackknife residual was used to diagnose the error (red dots). The scatter are the individual data points in the sample and are underlaid as a guide to aid manual verification.

LEFT PLOT: shows 3 exemplars had no accompanying decrease in WT (part C2a). The navy line shows a child with one decrease but two low measures, such that removing the first error meant another HT decrease remained, the screening routine for C2 was thus looped until no decreases remained.

RIGHT PLOT: shows 2 exemplars of HT decreases with a concomitant decrease in WT (part C2b), possibly due reflecting an incorrect date. Both HT and WT values were removed (red dots).

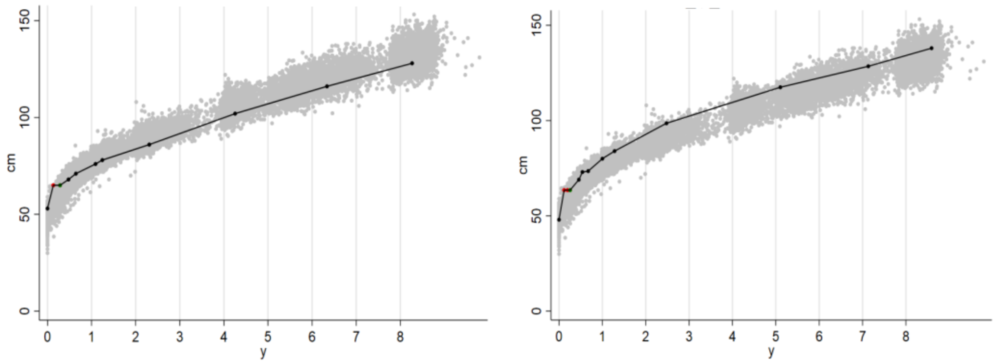


Figure S7 Exemplar plots of 2 children with duplicate HT values that manifested as implausible HT increases and were flagged in part C3a of the algorithm. The scatter are the individual data points in the sample and are underlaid as a guide to aid manual verification.

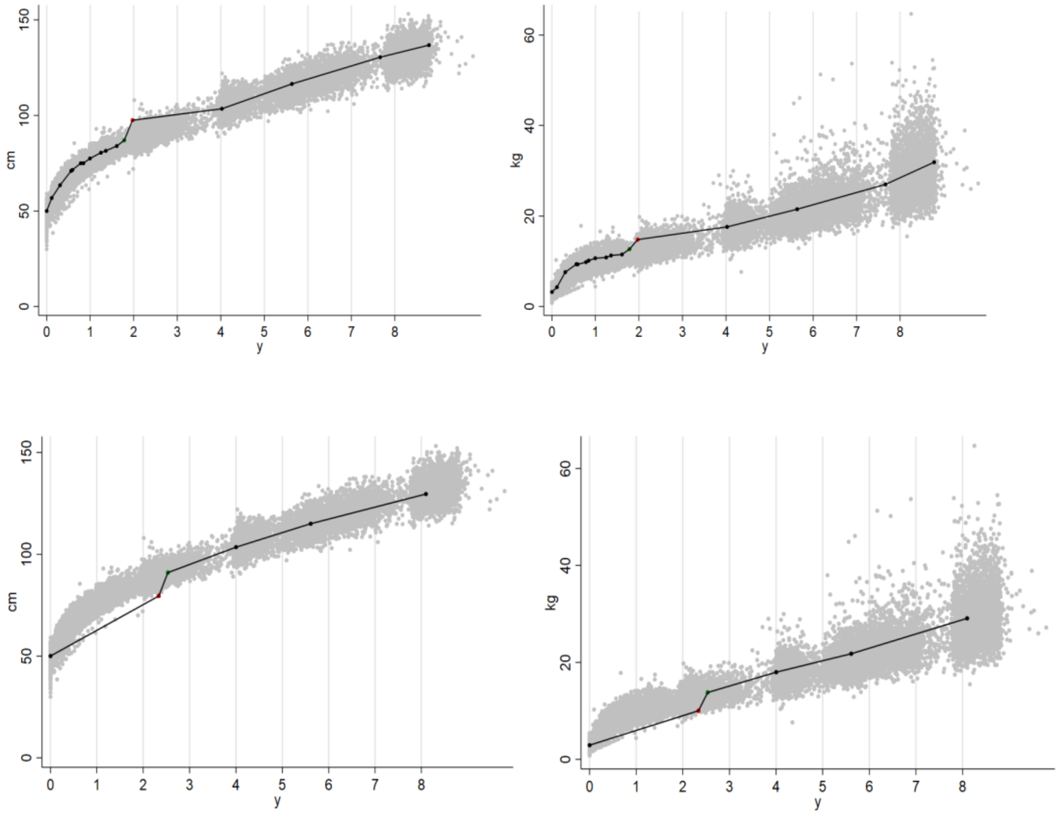


Figure S8. Exemplars of HT and WT growth histories from two children (top row & bottom row) with data that were flagged as impossible increases in HT (part C3b and C3c). In both children the impossible HT increase also occurred alongside concomitant WT increases, which may reflect a date error. The red value was diagnosed as the error according to the automated routine which selected the most internally inconsistent value. The scatter are the individual data points in the sample and are underlaid as a guide to aid manual verification.

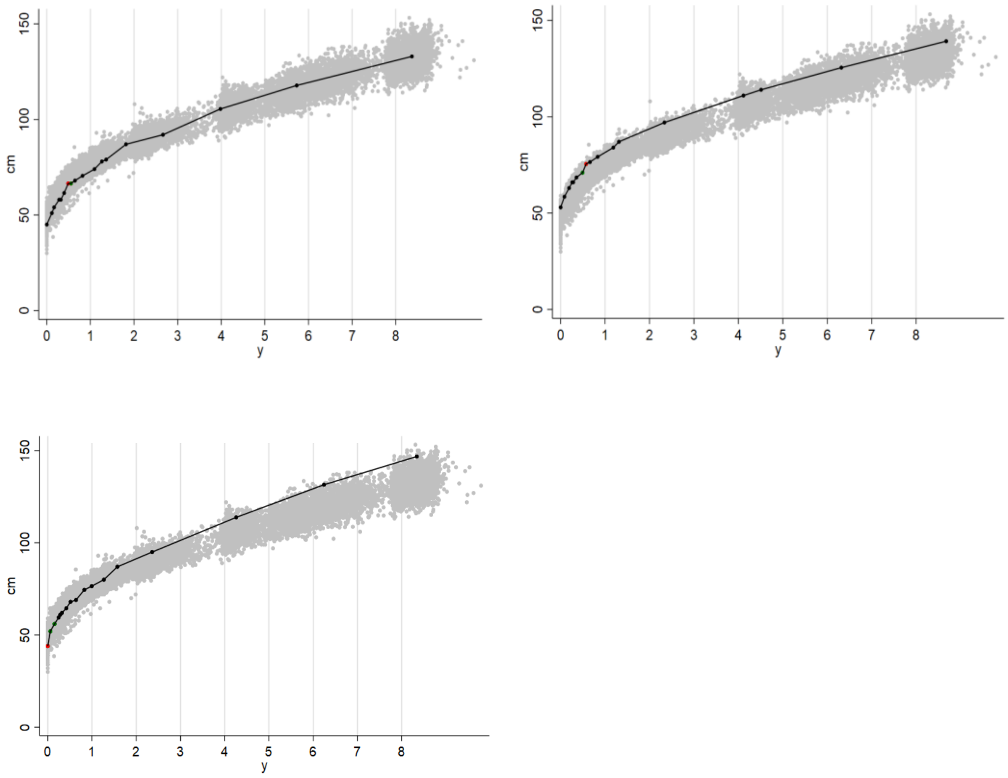


Figure S9. Exemplars from three children with flagged values exceeding the velocity threshold for HT increase (part C3c) but where the automated decision was either over-ridden or the value was kept because of plausible catch-up growth. The scatter are the individual data points in the sample and are underlaid as a guide to aid manual verification.

TOP PLOTS: Two exemplars where HT velocity exceeded the threshold but values were kept. Both originated from pairs of HT values close in time, where minor changes in HT within the expected accuracy of the measurement device can cause noisy first derivative (velocity) values.

BOTTOM PLOT: An exemplar of pre-term catch up growth in a child born at 34 weeks gestational age. This value exceeded the velocity threshold but was not removed as it is expected among pre-terms.

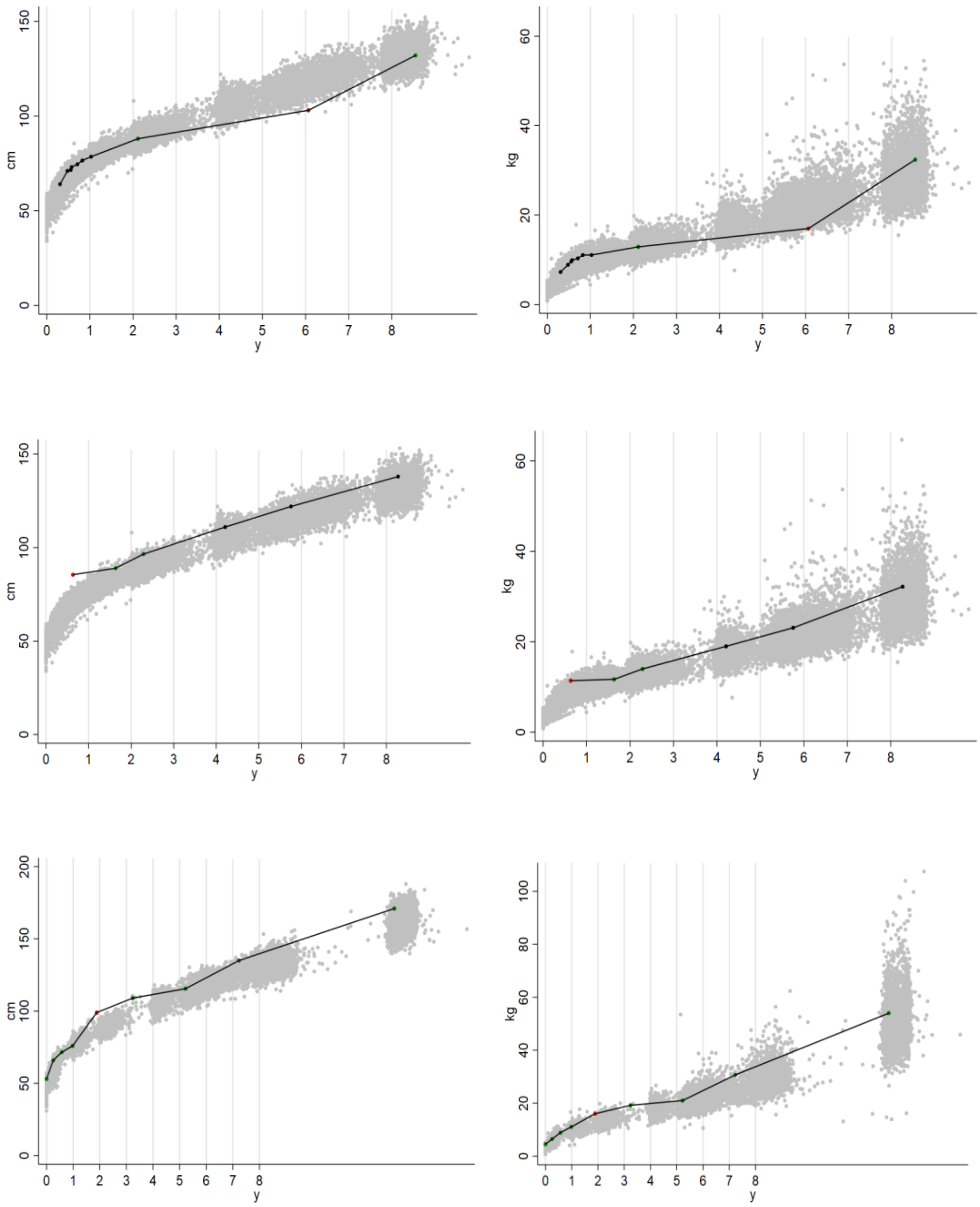


Figure S10. Exemplars of children flagged with inconsistent HT & WT patterns (part C4a). The scatter are the individual data points in the sample and are underlaid as a guide to aid manual verification.

TOP & MIDDLE PLOTS: show the HT and WTs of 2 children (one child per row). The red value was diagnosed as the error according to the automated routine which selected the most internally inconsistent value, and it was removed for both HT and WT.

BOTTOM PLOT: shows the HT and WT from a child that was flagged with implausible patterns (part C4a) but was over-ridden and kept after manual verification because of uncertainty over the genuineness of the value.

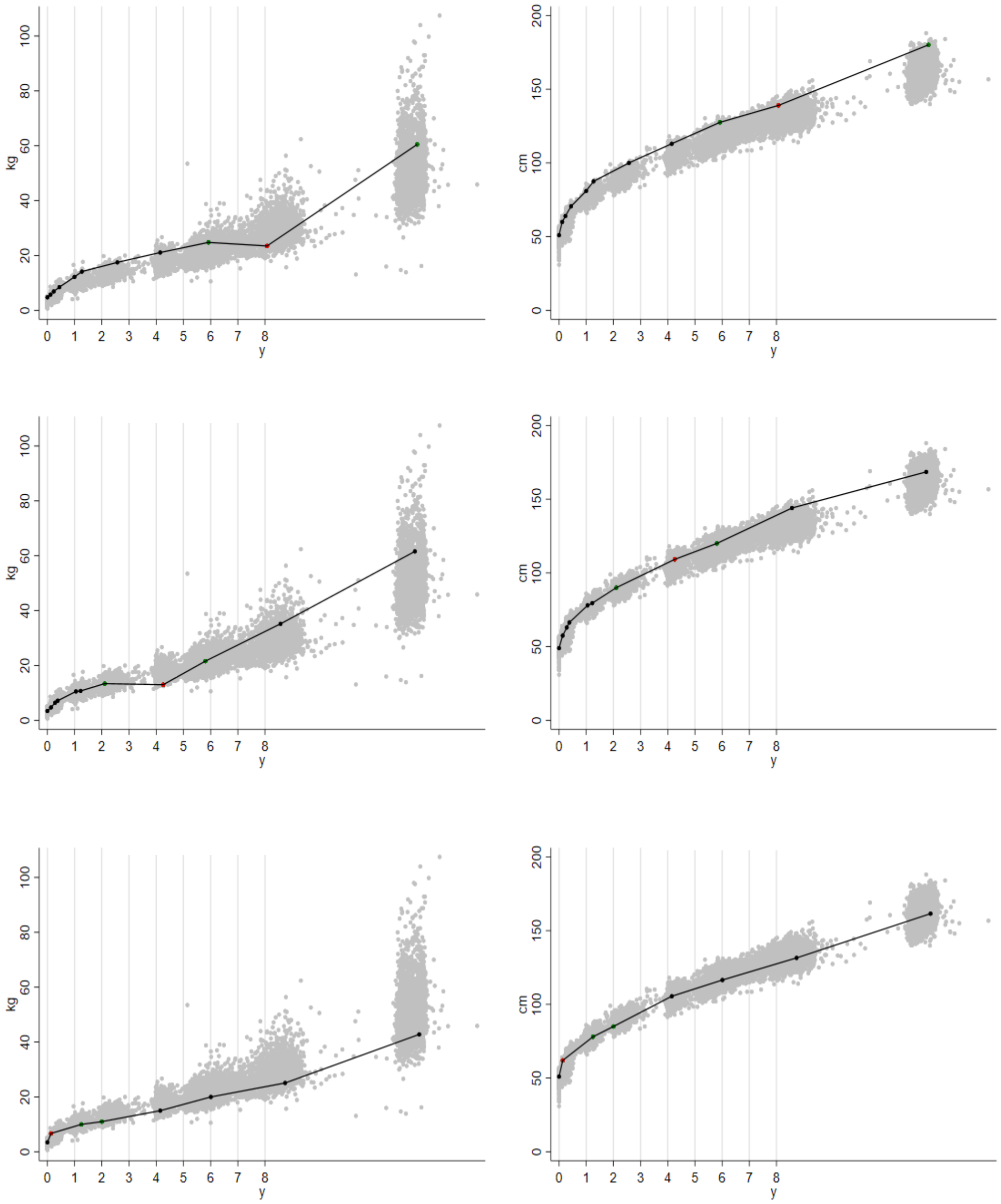


Figure S11.  Exemplars of inconsistent WT pattern decreases that were flagged in part C5 of the algorithm. The scatter are the individual data points in the sample and are underlaid as a guide to aid manual verification. Each row shows the HT and WT histories from the same individual

TOP & MIDDLE ROW: Both these exemplars show as a stalling in growth which is captured as a decrease on the age and sex standardised SD scale.

BOTTOM ROW: An exemplar of a WT stalling early in life that was over-ruled with the flagged value kept.

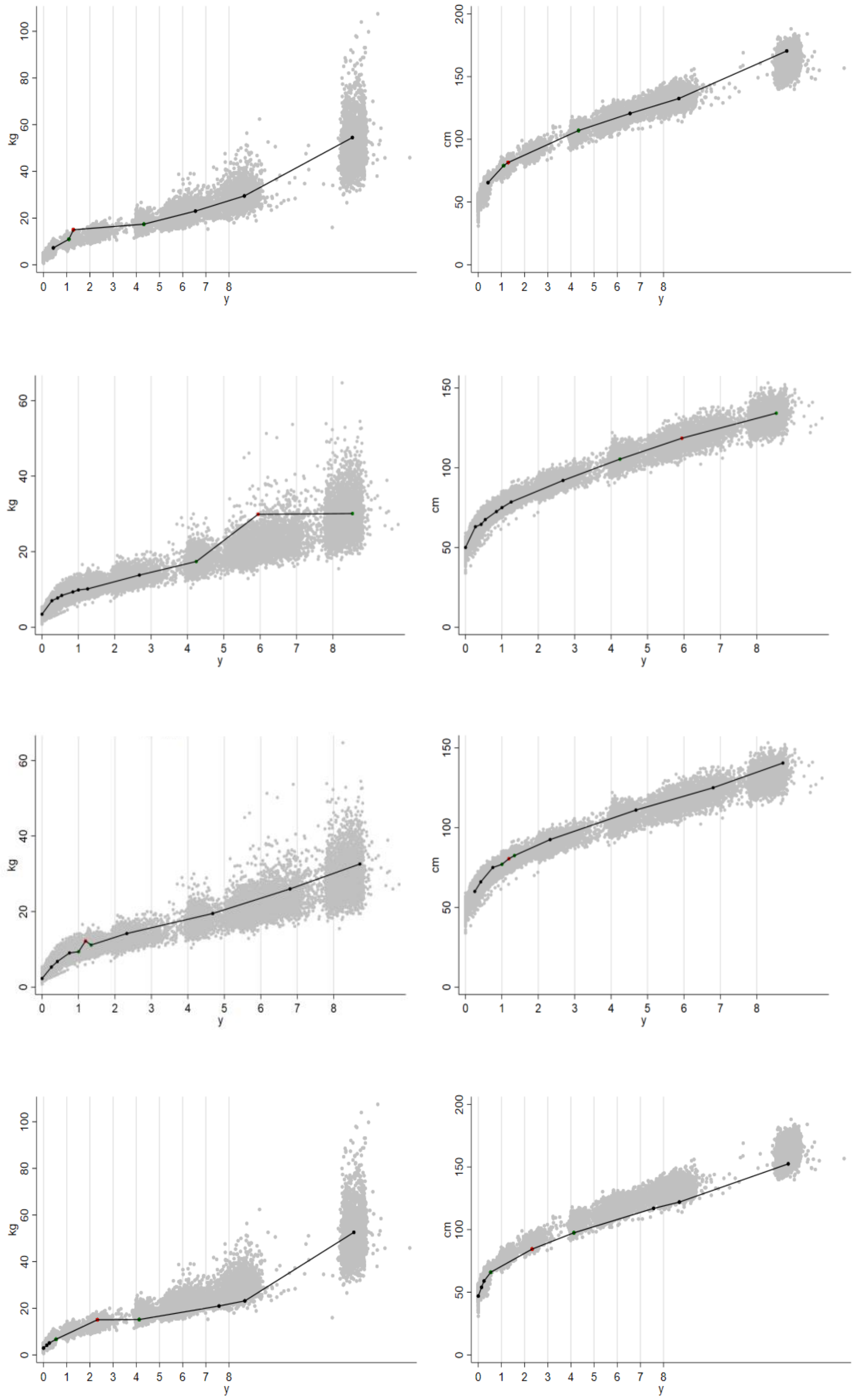

Figure S12. Exemplars of inconsistent WT pattern increases that were flagged in part C6 of the algorithm. The scatter are the individual data points in the sample and are underlaid as a guide to aid manual verification. Each row shows the HT and WT histories from the same individual

TOP 3 ROWS: Three exemplars of a WT pattern increase, the automated decision for the error is shown in red, these WT values (not HT) were removed.

BOTTOM ROW: An exemplar of a WT pattern increase that was over-ruled such that all values were kept.

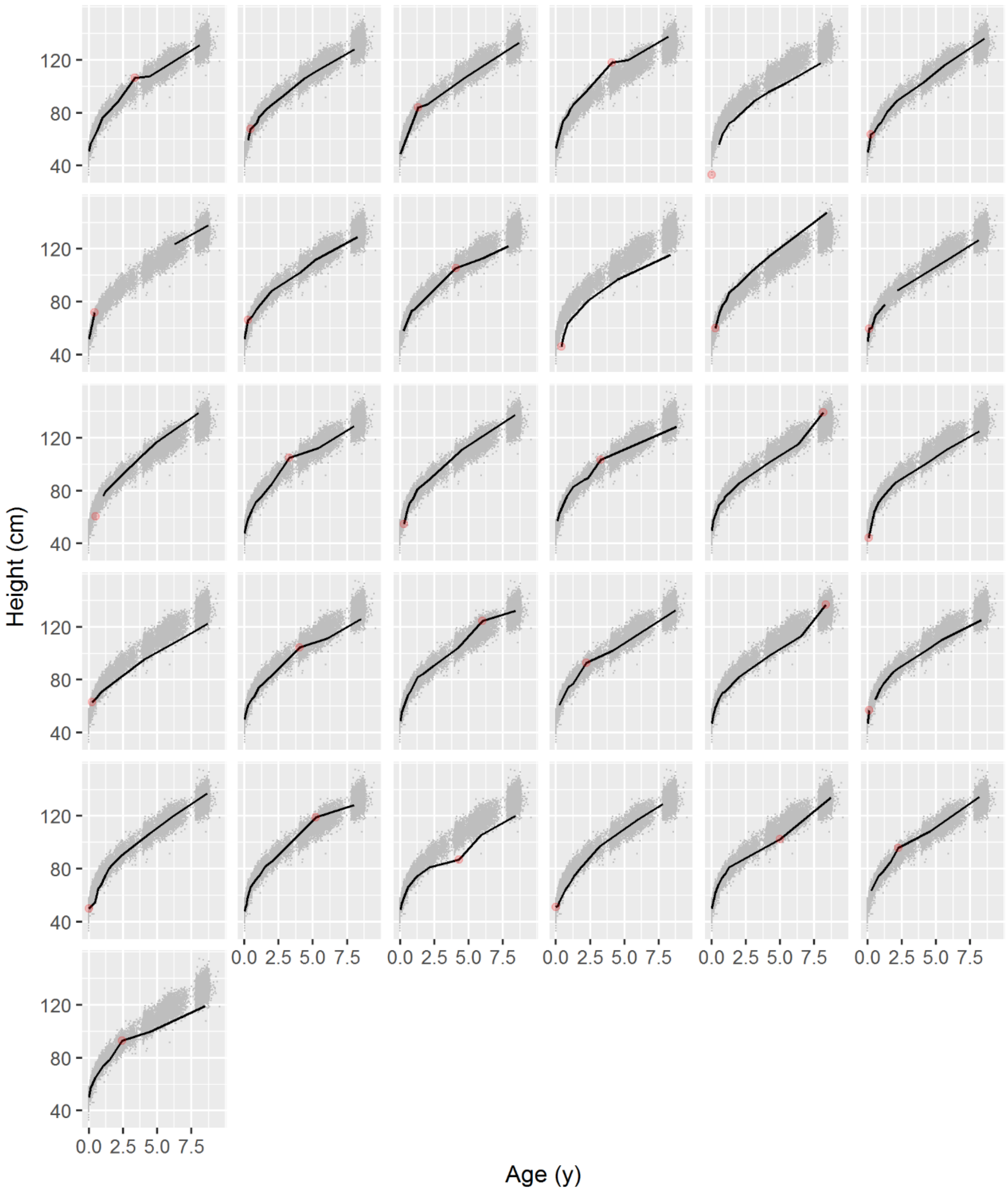


Figure S13. Spaghetti plots of HT from the 33 children from the 2010 dataset who were flagged in the Daymont algorithm (ref) as containing an suspicious value/ error (red dot) after the final run of the cleaning algorithm. None of these were considered implausible/ impossible without some uncertainty and so were kept.

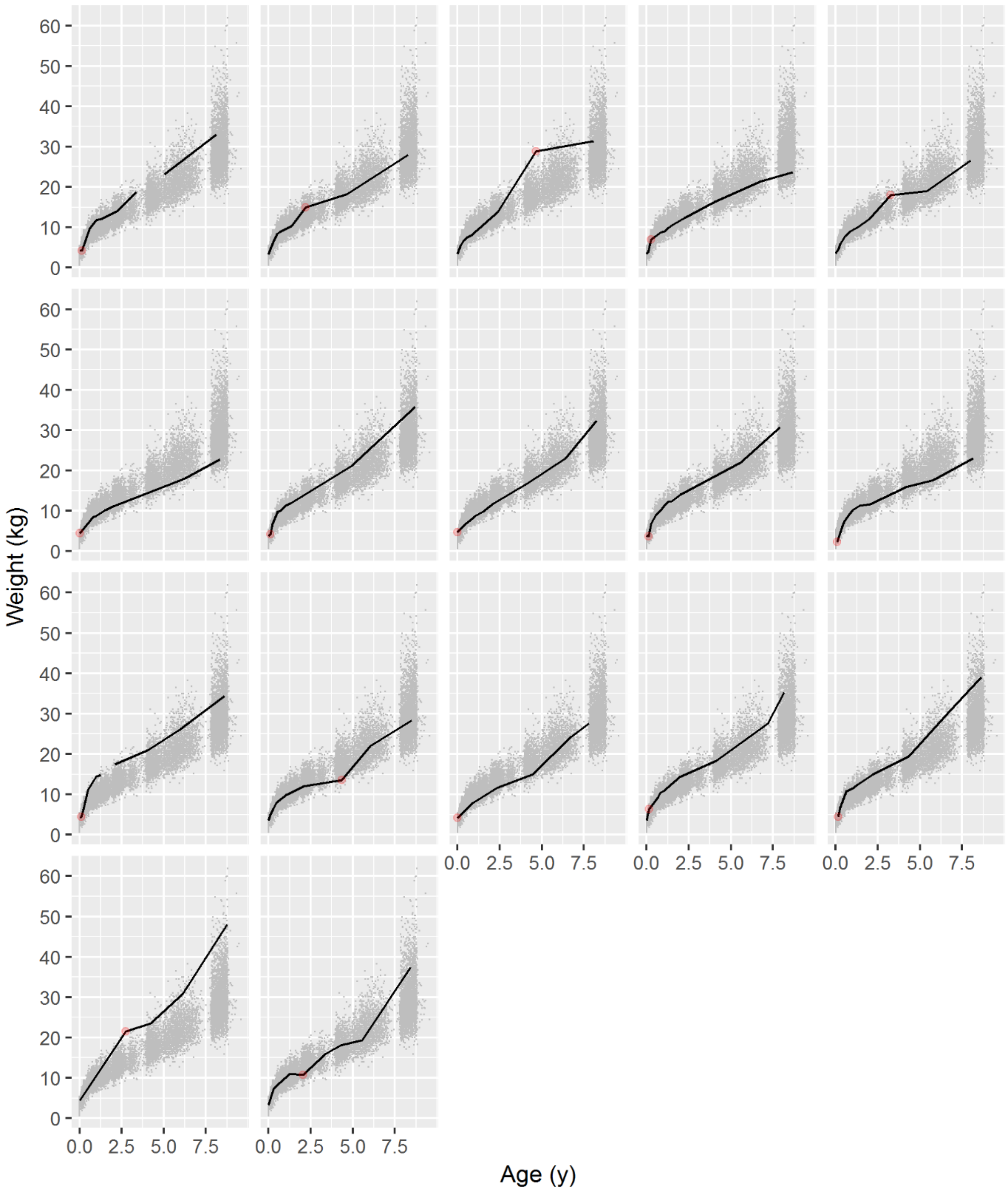


Figure S14. Spaghetti plots of WT from the 17 children from the 2010 dataset who were flagged in the Daymont algorithm as containing an error (red dot) after the final run of the cleaning algorithm. None of these were considered implausible/ impossible without some uncertainty and so were kept.

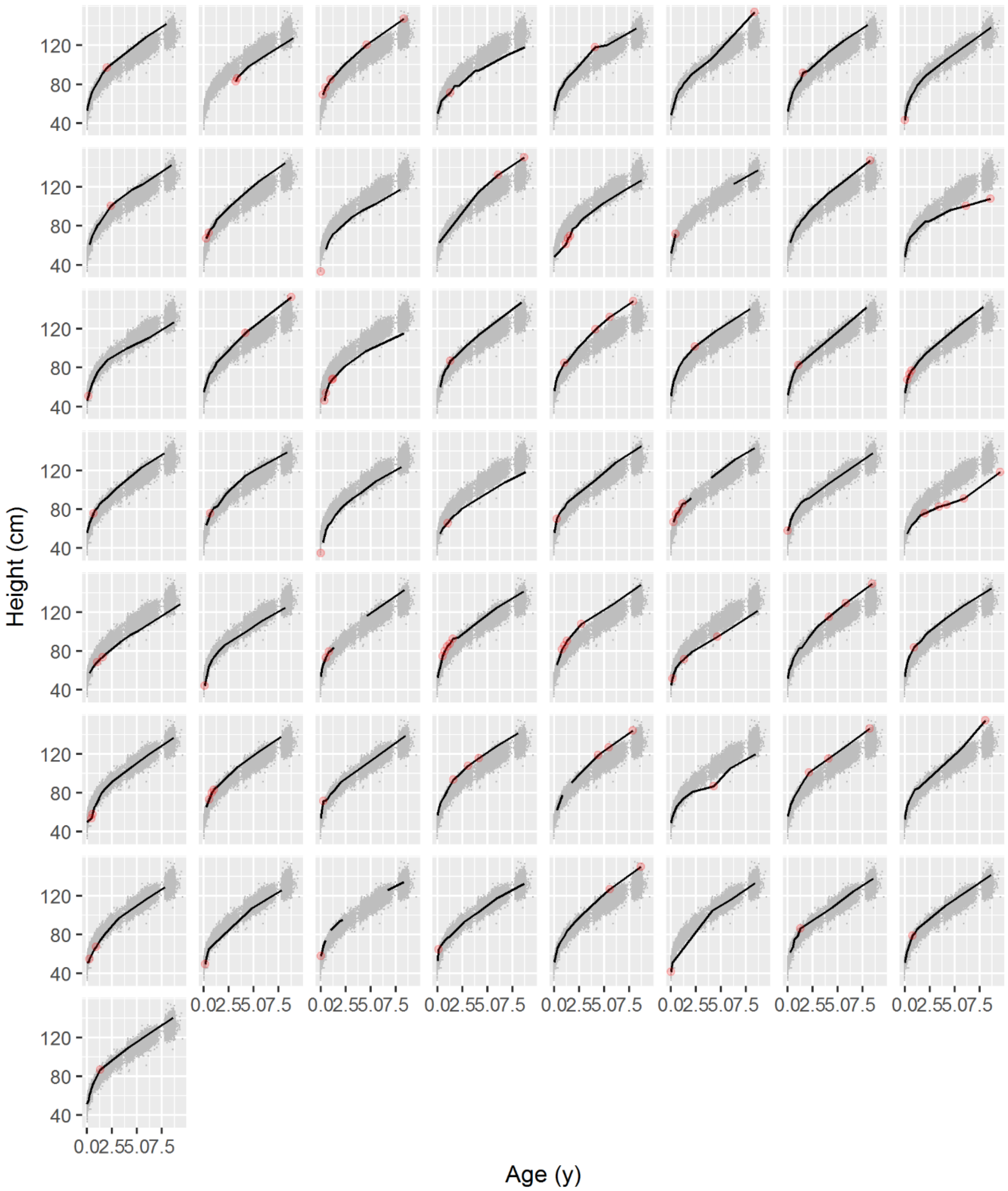


Figure S15. Spaghetti plots of HT from the 57 children from the 2010 dataset who at least one high or low modified CDC SD scores (|SD|>3) (red dot) after the final run of the cleaning algorithm. None of these were considered implausible/ impossible without some uncertainty and so were kept.

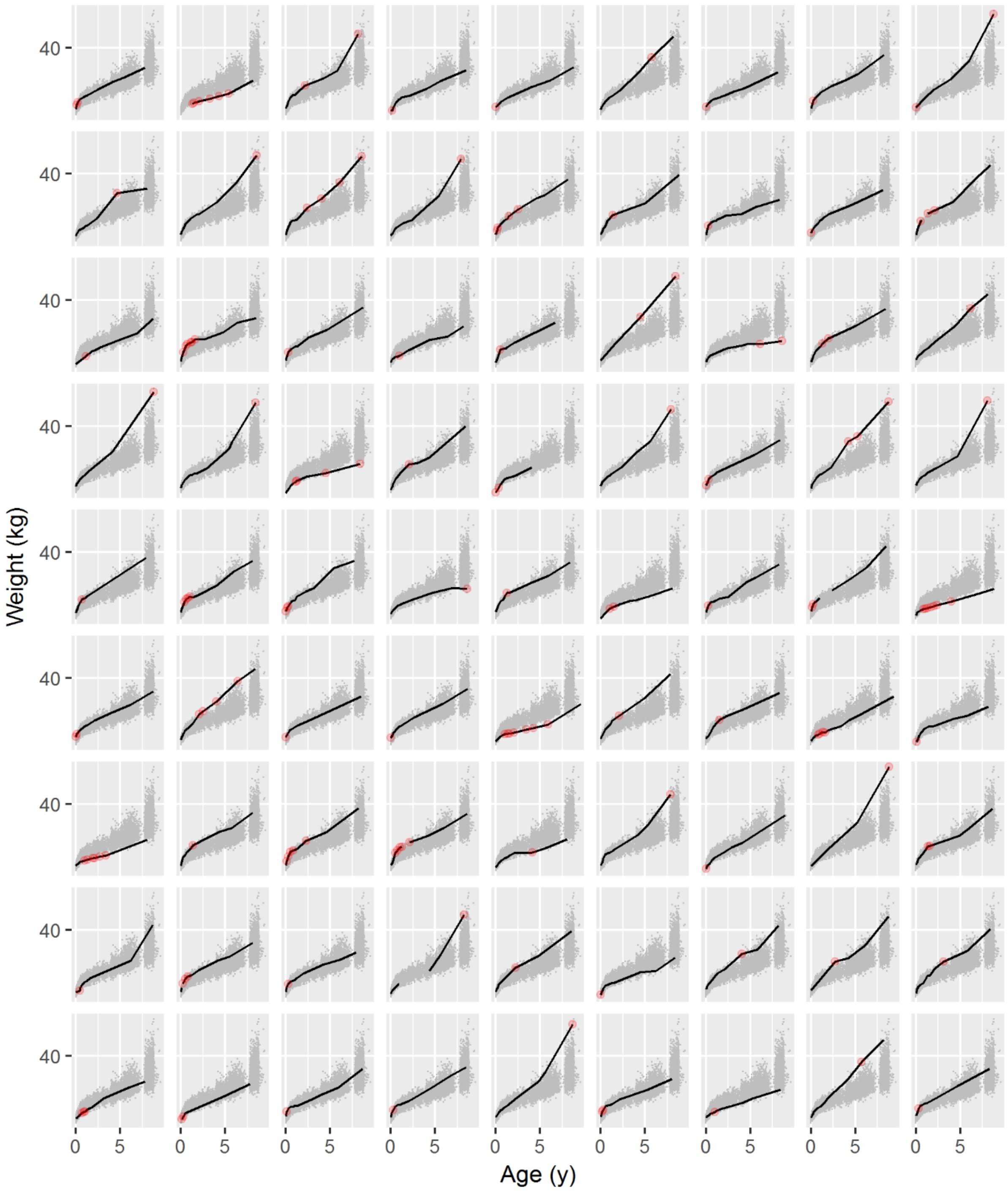


Figure S16. Spaghetti plots of WT from the 81 children from the 2010 dataset who at least one high or low modified CDC SD scores (|SD|>3) (red dot) after the final run of the cleaning algorithm. None of these were considered implausible/ impossible without some uncertainty and so were kept.
